# Finding Long-COVID: Temporal Topic Modeling of Electronic Health Records from the N3C and RECOVER Programs

**DOI:** 10.1101/2023.09.11.23295259

**Authors:** Shawn T. O’Neil, Charisse Madlock-Brown, Kenneth J. Wilkins, Brenda M. McGrath, Hannah E. Davis, Gina S. Assaf, Hannah Wei, Parya Zareie, Evan T. French, Johanna Loomba, Julie A. McMurry, Andrea Zhou, Christopher G. Chute, Richard A. Moffitt, Emily R Pfaff, Yun Jae Yoo, Peter Leese, Robert F. Chew, Michael Lieberman, Melissa A. Haendel, the N3C Consortium and the RECOVER Consortium

## Abstract

Post-Acute Sequelae of SARS-CoV-2 infection (PASC), also known as Long-COVID, encompasses a variety of complex and varied outcomes following COVID-19 infection that are still poorly understood. We clustered over 600 million condition diagnoses from 14 million patients available through the National COVID Cohort Collaborative (N3C), generating hundreds of highly detailed clinical phenotypes. Assessing patient clinical trajectories using these clusters allowed us to identify individual conditions and phenotypes strongly increased after acute infection. We found many conditions increased in COVID-19 patients compared to controls, and using a novel method to associate patients with clusters over time, we additionally found phenotypes specific to patient sex, age, wave of infection, and PASC diagnosis status. While many of these results reflect known PASC symptoms, the resolution provided by this unprecedented data scale suggests avenues for improved diagnostics and mechanistic understanding of this multifaceted disease.

## Introduction

The long-term health consequences of SARS-CoV-2 are not fully understood.^1^ Research suggests that many patients experience persistent symptoms,^2–4^ known as Post-Acute Sequelae of SARS-CoV-2 infection (PASC; also known as Long-COVID), affecting multiple organ systems, including pulmonary, cardiovascular, hematological, neurological, and renal systems.^5^ The disruption of the host immune response is suspected to play a role in various PASC-associated conditions, including reactivation of dormant persistent infections,^6^ autoimmune responses,^7^ and multi-inflammatory syndrome of children (MIS-C).^8^ Emerging evidence indicates that PASC may present with one or more sub-phenotypes, comprising potentially overlapping clusters of frequently co-occurring symptoms, and prevalence of these may be influenced by patient demographics or infection severity.^4,9–12^

Various clustering algorithms have been applied to identify sub-phenotypes of PASC from patient data, including k-means clustering with disease ontology data, multiple correspondence analysis, hierarchical ascendant classification, association rule mining, and latent class analysis.^9,10,13–15^ Topic models, a class of natural-language clustering techniques suitable for electronic health record (EHR) data, are also widely used. Broadly, topic models identify ‘topics’ as sets of frequently co-occurring terms in a document corpus. In EHR contexts, a patient’s clinical history is treated as a document, and their conditions or other events serve as terms, often encoded in medical vocabularies such as ICD-10-CM codes. Topic modeling methods employed for PASC and COVID-19 include Poisson factor analysis, non-negative matrix factorization, and Latent Dirichlet Allocation (LDA).^16–18^ LDA is particularly well studied in the context of EHR data,^19,20^ characterizing each topic (interpreted as a phenotype or sub-phenotype depending on context) as a distinct probability distribution over terms (conditions or other medical events), and each document (patient) as a distinct distribution over topics.

While these studies have identified PASC sub-phenotypes in unique ways, common themes have emerged, including clusters representing cardiovascular,^9,10,16,18,21^ pulmonary,^10,14,16,21^, neurocognitive,^10,14^, musculoskeletal,^9,10,14,16,21,22^, and fatigue-related symptoms.^10,14,21^ In many cases these are combined and there exist one or more multi-system clusters.^9,10,13,15^ Despite the diverse methods applied, a variety of limitations prevent understanding these sub-phenotypes in a broader clinical context. For example, many studies cluster symptoms and diseases from the post-infection period only,^9,10,13–17,22^ and do not consider the presence of pre-existing sub-phenotypes beyond predetermined risk factors (though see Humpherys et al. who investigate mortality risk via independent pre- and post-infection clusterings^18^). Though some studies focus only on patients suspected or diagnosed with PASC,^9,10,13,16^ or differentiate between COVID-19 patients with and without PASC symptoms,^14,21^ few include individuals lacking any indicators of COVID-19.^15,22^ Without this broader comparison, identifying sub-phenotypes unique to these diseases is challenging. Finally, while clustering methods are naturally data driven, some research limits scope to conditions already presumed linked to PASC,^9,13,15,16^ potentially missing rare or otherwise unknown associations.

To better understand the long-term effects of COVID-19, we employed LDA topic modeling on EHR data from the National COVID Cohort Collaborative (N3C), encompassing over 12 million patients across 63 clinical sites and more than 230 healthcare locations.^23^ The resulting model identified hundreds of clinically-consistent condition clusters as topics representing potential sub-phenotypes of interest. New-onset rates for a diverse set of top-weighted conditions were significantly higher in held-out PASC and COVID-19 patients compared to Controls with no COVID-19 indication. Further, by analyzing whole-topic associations temporally (pre- and post-infection) among these cohorts, we identified several sub-phenotypes associated with COVID-19 or PASC, specific to patient age, sex, or pandemic wave. Many of our findings confirm established features of PASC, while others present new insights or more detailed perspectives, prompting further investigation into this complex and heterogeneous condition.

## Methods

### Study design

The LDA implementation we use aims to represent a document corpus as a set of *topics*, where each topic is characterized by a multinomial distribution over possible words or terms, and each document is characterized by a multinomial distribution over topics. It assumes a hierarchical generative model, where for any given term in a given document, a topic is first selected according to the document’s topic distribution, and then a term is selected according to that topic’s term distribution.^24^ In our application patient medical histories represent documents and conditions recorded for those patients represent terms, a common approach in topic modeling for EHR data.^19,20^

Figure 1 illustrates our approach. Initially, we trained a model on a diverse set of patient histories, including those with and without COVID-19, using independent training and validation patient sets to guide model development. The resulting topics represent a comprehensive set of phenotypes across the N3C cohort; to understand how these topics relate to patients over time and with respect to infection status, we held out an additional independent *assessment* patient set. Within this set we identified three cohorts: PASC, patients with a clinical PASC diagnosis, COVID, those with an indicated COVID-19 infection but no PASC diagnosis, and Control, those with neither. These patients’ histories were divided into pre- and post-infection clinical phases, assigning Control patients a mock infection date for the purpose. In a first set of statistical tests we individually assessed the top 20 high-relevance conditions from each topic, comparing new-onset rates post-infection for PASC and COVID patients compared to Controls. Next we assessed each topic as a whole, by modeling topic probability estimates derived separately from pre- and post-infection data. Including patient demographics such as age and sex in these models reveals which patient cohorts and groups experience significant changes in which topics post-infection, as compared to Controls.

**Figure 1:**
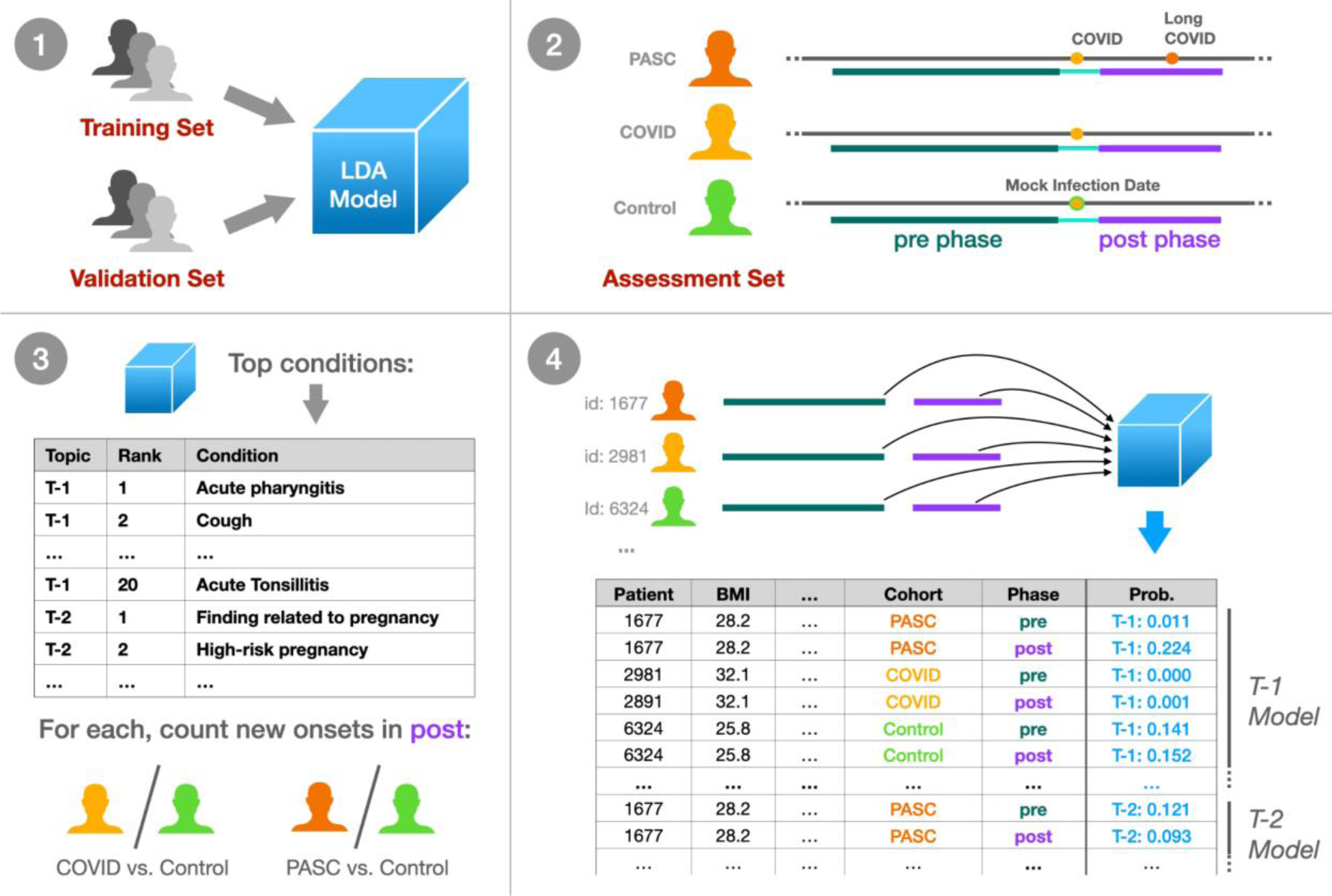
Experimental design summary. (1) We trained an LDA topic model on a broad set of N3C patient data, tuning and evaluating the model with a held-out validation set using the UCI coherence metric. (2) Within a separate held-out assessment patient set, we defined three cohorts: PASC (patients with Long COVID), COVID (COVID-19 only), and Control (neither). For these patients we defined a 1-year pre-infection phase 6-month post-infection phase, utilizing a mock infection date for Control patients. (3) For the top 20 conditions per topic, we assessed new onset rates for COVID and PASC patients compared to Controls in the post-infection phase. (4) Finally, we defined per-topic logistic models, with outcome variables as the topic model’s assigned probabilities to individual patient phase data. Model coefficients then relate patient demographics, cohort, infection phase, and combinations of these factors to topic assignment for further study.

### Data pre-processing

N3C multi-site EHR data are harmonized to the OMOP common data model. We analyzed condition records from OMOP’s condition_era table, which merges repeated, identical condition records into single condition timeframes via a 30-day sliding window.^25^ We used N3C release v87, representing data as of Aug. 2, 2022, using only records from a subset of sites passing minimal quality filters (see Suppl. Methods). The COVID-19 diagnostic code (U07.1), being a major criterion for N3C data selection and present in 22% of patients, was excluded, along with early pandemic alternatives such as *unspecified viral disease* (10%) and *disease due to coronaviridae* (0.5%). Additional clinically-uninformative terms such as *Clinical finding* and *Findings of sexual activity* were removed, as were all entries not expected in the OMOP condition domain (Suppl. Methods, Suppl. Table 1). Records with implausible dates, either starting before January 1, 2018 (N3C’s earliest inclusion date) or extending beyond a site’s data contribution date, were also excluded. Patients were randomly assigned to sets, with 20% allocated to the assessment set and the remaining patients split 80% for training and 20% for validation (Suppl. Figure 1).

### Cohort selection and clinical phases

Before conducting statistical analyses, we filtered patients in the assessment set to ensure data quality and consistency. Some N3C-contributing sites reported no U09.9 PASC diagnosis codes, possibly due to lack of implementation in their EHR software,^26^ leading us to exclude all patients from these sites to prevent misclassification of PASC patients. For individual-condition tests, we required patients to have at least two weeks of active condition history in both pre- and post-infection phases. For per-topic tests, we assessed patient features requiring complete data across all covariates and excluded Omicron patients due to incomplete data (see Statistical analyses below).

Each patient in the assessment cohorts was assigned an index date, representing their estimated first COVID-19 infection, or a mock infection date for Control patients. From these dates we identified two clinical phases per patient: a 1-year, pre-infection phase ending 15 days before the index (to account for acute symptoms prior to first diagnosis or test), and a 6-month, post-infection phase beginning 45 days after the index to capture possible PASC conditions.^27^ Conditions were counted in any phase they overlapped, allowing a single condition to be represented in both phases if applicable. Cohorts were restricted to patients supporting these three contiguous phases within a single observation period, as recorded in the OMOP observation_period table.

### PASC

Patients with a U09.9 PASC diagnosis code on or after Oct. 1, 2021 when this code was released, or the CDC-recommended alternative B94.8 *Sequelae of other specified infectious and parasitic diseases* prior to this date.^28^ For patients with a strong primary infection indicator, a positive SARS-CoV-2 Polymerase Chain Reaction (PCR) or Antigen (Ag) test (Suppl. Table 2) or U07.1 COVID-19 diagnosis, we chose the first of any of these as the infection index date. For patients without these we used the first PASC indicator as the index date. In cases where the primary infection indicator occurs within 45 days of PASC we considered the test or diagnosis unreliable and used the PASC indicator as the index date.

### COVID2

Patients with a confirmed primary COVID-19 infection as indicated by a positive PCR test, antigen test, or U07.1 COVID-19 diagnosis, and who are not in the PASC cohort. Their infection index date was the first of any of these indicators.

### Control

Patients with no indication of COVID-19 in N3C data, including positive PCR, antigen, or antibody test (Suppl. Table 3), COVID-19 (U07.1) or PASC diagnosis (U09.9 or B94.8), or a visit to a PASC specialty clinic (unique information provided by six N3C-contributing sites). Patients diagnosed with *Multisystem inflammatory syndrome* (M35.81) were excluded as potential confounders. Control patients were assigned a mock infection date, chosen uniformly at random to simulate pre-infection, acute, and post-infection phases of the correct lengths contained entirely within their longest continuous observation period. Mock infection dates were constrained to be after March 1, 2020 to align temporally with pandemic trends.

### Machine learning methods

To train our topic model, we used full pre-processed N3C patient histories from the training set as documents, employing per-patient counts of OMOP condition_concept_id entries from the condition_era table. While many LDA variations exist, most are computationally prohibitive for the scale of data considered. We therefore adopted the online LDA method described by Hoffman et al.,^24^ implemented in Apache Spark version 3.2.1,^29^ with a 5% batch size and 10 iterations over the data (maxIter = 200, subsamplingRate = 0.05).

Using the held-out validation patient set we computed UCI Coherence^30^ to measure model quality and choose the final number of topics. For each topic, this unitless metric assesses how frequently top-weighted conditions co-occur in patients relative to random chance (see Suppl. Methods), with higher values indicating higher-quality topics. As coherence scores are normally distributed (see Results), we report coherence as a z-score *C,* with positive values indicating higher-than-average topic coherence and quality.

We further defined a usage value *U* (range 0–100%) as the average assigned probability across patients. Given that topic usage varied across N3C-contributing sites, we calculated a usage uniformity metric *H*, expressed as the normalized information entropy (range from 0–1) of site usage, with values approaching 1.0 indicating more uniform usage across sites. We also conducted topic similarity analysis via Jensen-Shannon distance (range 0–1), a symmetrical metric with values closer to 0.0 suggesting more similar topics.^19,31,32^

Common conditions such as *Essential Hypertension* may be highly weighted by many topics. A condition’s *relevance* to a topic is the log-ratio of the topic-specific probability to the condition’s global probability (as defined by the LDAVis package^33^ when γ = 0), with positive values indicating conditions more specific to a given topic. Highly-weighted terms with low relevance thus indicate those heavily used by multiple topics. Topic usage and term relevance were computed using both the validation and training sets for completeness.

### Statistical analyses

As discussed above, we used assessment cohort data to determine how individual conditions, and whole topics, uniquely manifest post-infection in PASC and COVID patients as compared to Controls.

Across topics, we selected the top 20 conditions with positive relevance scores for individual testing of new-onset rates, a count chosen to balance the total tests required with depth of topic exploration, and roughly aligned with our topic visualizations (see Results). For each condition we considered patients with no incidence in the pre-infection phase, counting those who did and did not go on to experience the condition in the post-infection phase per cohort. 2×2 Fisher’s exact tests assessed these counts for PASC versus Control and COVID versus Control patients separately. Tests were multiply-corrected (Bonferroni) and used fisher.test in R (v3.5.1) with `alternative` `=` `“two.sided”` and simulate.p.value = TRUE to allow for large and small counts.^34^

For whole-topic assessment, we used the trained topic model to independently estimate posterior topic distributions for pre- and post-infection data. Given the generative model assumed by LDA, a probability of *p* for topic T in phase *i* suggests that *p*% of newly sampled conditions in *i* would be sourced from T.^24^ More formally, single-topic probability estimates follow a Beta distribution as a result of the Dirichlet prior.^35^ For each topic we model these as success rates in binomial trials (Figure 1), resulting in a potentially overdispersed Beta-binomial distribution.^36^ We thus use Generalized Estimating Equations (GEEs; geeglm in geepack v1.3.9), both to capture within patient pre- and post-infection correlation structure (with id = person_id, corstr = “exchangeable”),^37^ and to employ robust error estimation while allowing for overdispersion (with scale.fix = FALSE).^38,39^ Outcomes were equally weighted, as opposed to weighted by the number of conditions represented, to avoid high-utilization patients dominating results.

In addition to phase (pre- or post-infection) and cohort (PASC, COVID, Control), each topic’s logistic model included patient demographic covariates: sex (Male, Female), race (White, Black or African American, Asian or Pacific Islander, Native Hawaiian or Other Pacific Islander, Other or Unknown), BMI, life stage (Pediatric 0-10, Adolescent 11-18, Adult 19-65, Senior 66+), Quan-based Charleson comorbidity index (Suppl. Methods),^40^ and date-based “wave” of infection. We defined infection waves based on CDC surveillance data,^41^ categorizing them as Early (prior to March 1, 2021), Alpha (March 1, 2021 to June 30, 2021), or Delta (July 1, 2021 to Dec. 31, 2021). Patients with index during the Omicron wave (Jan. 1, 2022 and later) were excluded due to limited data across covariates, as were all patients without complete information. Models included site-level covariates as potential sources of heterogeneity,^28,42^ including source common data model (PCORnet, ACT, OMOP, TrinetX, OMOP-PedsNet), percentage of PASC patients, and site-specific topic usage (Suppl. Methods). We also developed models without site-level covariates for a subset of topics to assess their importance.

After fitting these models, we applied a difference-in-differences approach using estimated marginal means contrasts to look for changes in topic rates pre-to-post infection, for PASC patients versus Controls, within specific groups defined by sex, life stage, and wave of infection. The same tests were run for COVID versus Control patients. To validate this approach, we conducted additional ‘baseline’ contrasts for expected differences in females versus males, and pediatric, adolescent, and senior patients versus adults. In total we conducted 22 contrasts for each topic, multiply-correcting the complete set across all topics (Holm). Estimated marginal means contrasts were provided by emmeans (v1.8.9) in R (v3.5.1).

## Results

### Topic usage and coherence across sites

Of 75 available sites, 63 passed initial quality filtering, representing 12,486,133 patients with at least one condition recorded between 1/1/2018 and 8/2/2022. The topic model training set contained 7,992,339 patients and 387,401,304 conditions, while the validation set contained 1,996,380 patients and 96,738,753 conditions, representing a corpus of 48,372 unique condition identifiers. Mean topic coherence improved as the number of generated topics increased from 150 to 300, but not beyond (Suppl. Figure 2), so we selected the model with 300 topics for final analysis.

Figure 2 illustrates selected topics as word clouds, displaying the top conditions of each by weight. Topics are named T-1 to T-300 in order of their usage *U* (rounded to nearest 0.1%, see Methods), font size is proportional to condition weight in each topic, and color indicates condition relevance to the topic. Supplementary materials include word clouds for all topics (Suppl. Figure 3). Jensen-Shannon distance indicates that topics have little overlap (Suppl. Figure 4), with a median distance of 0.82 (range 0.39–0.83). The last 10 topics however, T-290 to T-300, form a group with increased co-similarity and many generic, low-relevance conditions mixed with a small number of high-relevance conditions.

**Figure 2:**
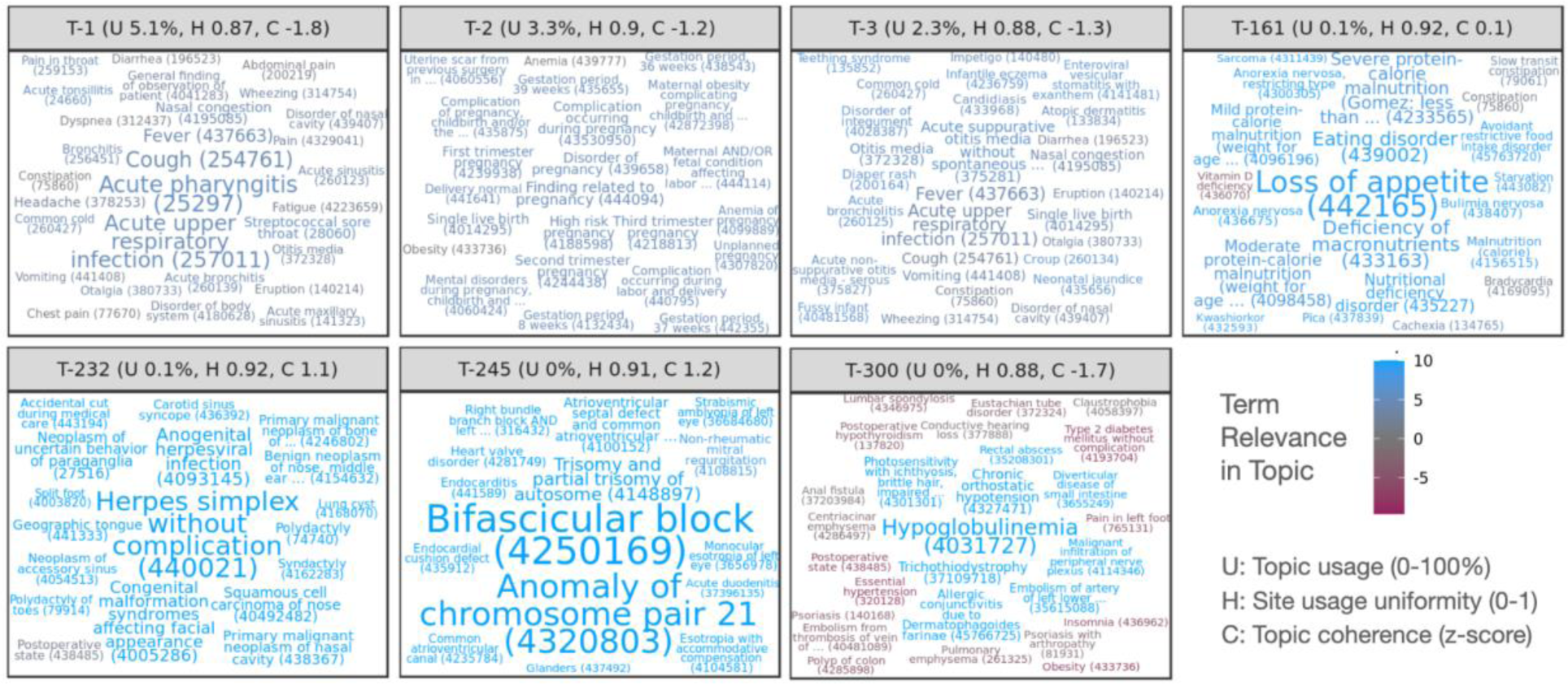
Word clouds illustrating top-weighted conditions for selected topics. Conditions are sized according to probability within each topic and colored according to relevance, with positive relevance indicating conditions more probable in the topic than overall. Each condition displays the numeric OMOP concept ID encoding the relevant medical code used for clustering, as well as the first few words of the condition name. Per-topic statistics in panel headers show usage of each of each topic across sites (U, rounded to nearest 0.1%), topic uniformity across sites (H, 0–1, higher values being more uniform), and relative topic quality as a normalized coherence score (C, z-score, higher values being more coherent).

Coherence scores follow a roughly normal distribution across topics (Suppl. Figure 5), and overall coherence tends to increase with rarer, more specific topics except for the last 10. Topic coherence varies by site, moreso for rarer topics (Suppl. Figure 6). All sites exhibit low coherence for the final 10 topics, and most of the final ∼35 are low coherence for most sites except for one. Two sites report low coherence for most topics. Topic usage also varies by site, though most sites and topics follow similar patterns of usage (Suppl. Figure 7). T-4 was used almost exclusively by a single site and has very low coherence with only a few high-relevance terms, although this site uses other topics similarly to other sites.

N3C sites contribute data from one of several source common data models (CDMs). The source CDM used by sites is not strongly correlated with coherence or usage (Suppl. Figures 6 and 7), except for two sites in the PEDSnet network specializing in pediatric care and another using TriNetX. These three sites exhibit distinctive patterns, including lower coherence and usage for T-153 pertaining to *Gout* (not typically associated with pediatric patients) and higher usage for T-127 pertaining to male pediatric conditions such as *Phimosis* and *Undescended testicle*.

### Individual conditions significant for PASC and COVID

From the 2,495,414-patient assessment set, 4,386 PASC, 105,967 COVID, and 335,841 Control patients met cohort eligibility requirements for individual-condition tests. Amongst PASC patients, 36% had a strong primary infection indicator at least 45 days prior to their PASC indication. After removing duplicates from the top entries for each topic, we tested 4,794 individual conditions for new onset post-infection. Of these, 213 are significant for the PASC cohort, 208 for COVID, and 89 for both with p < 0.05 after multiple correction. The complete list of significant results is available in Suppl. Table 4, and Figure 3 labels a subset of these. The PASC cohort shows larger rates for most significant conditions, although several conditions are represented in the COVID cohort as well, such as *Pneumonia caused by SARS-CoV-2*, *Viral pneumonia*, *Postviral fatigue syndrome*, *Loss of sense of smell*, and *Abnormal menstrual cycle.* Additionally, the following conditions have significant estimated odds ratios (ORs) greater than 2 in both cohorts: *Loss of sense of smell*, *Disorder of respiratory system*, *Acute lower respiratory tract infection*, *Upper respiratory tract infection due to Influenza*, *Telogen effluvium*, and *Non-scarring alopecia*.

**Figure 3:**
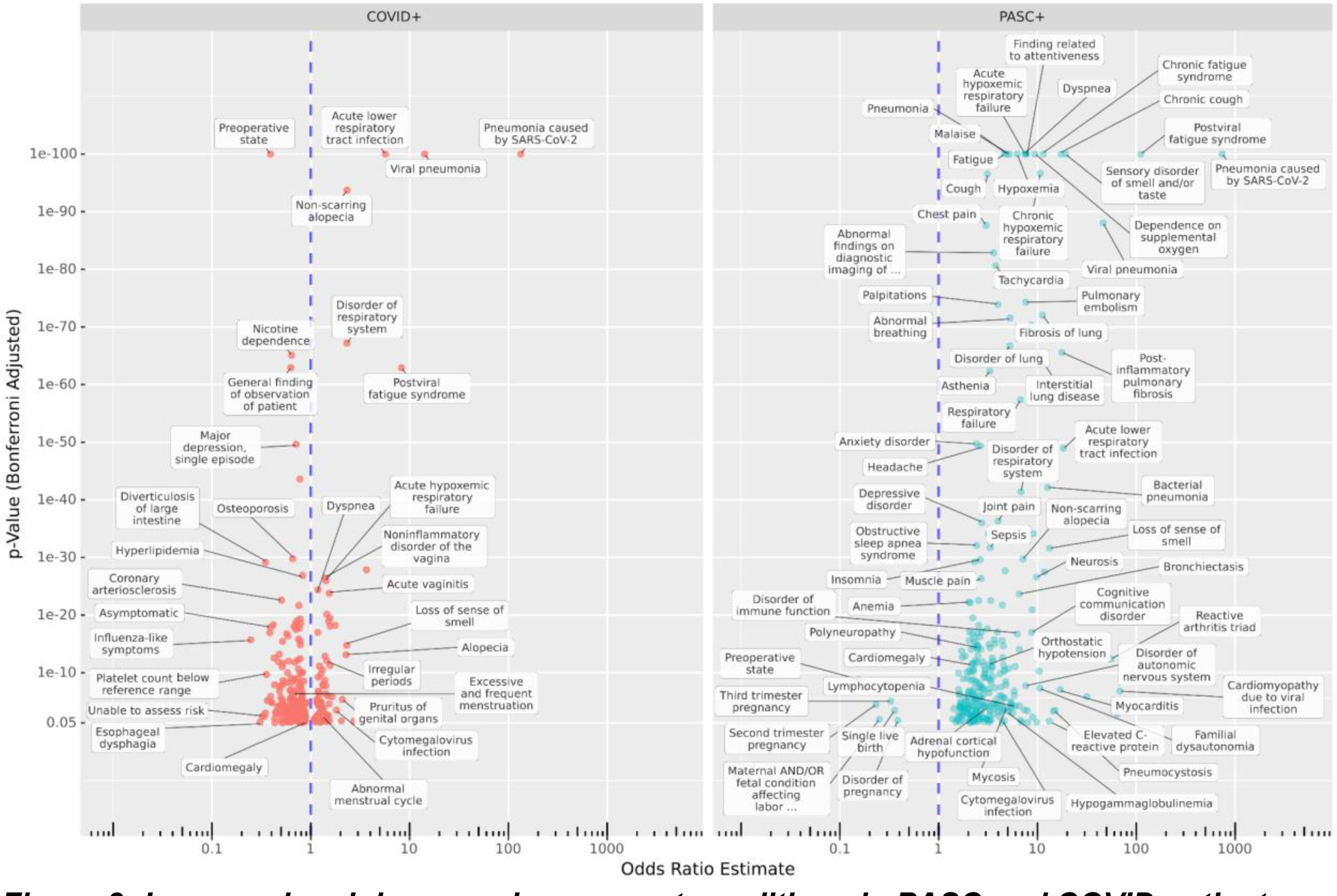
Increased and decreased new-onset conditions in PASC and COVID patients compared to Controls post-infection. The x-axis shows estimated odds ratios and the y-axis shows the adjusted p-values for new incidence of top-weighted, positive-relevance terms from all topics amongst COVID (left) and PASC (right) cohorts compared to Controls, in the six-month post-acute period compared to the previous year. Many known PASC-associated conditions increased in both cohorts, while some conditions are cohort-specific. Additionally, in the COVID cohort, incidence of many conditions associated with regular care or screening is reduced compared to controls.

Several conditions are strongly increased in the PASC cohort, including *Chronic fatigue syndrome*, *Malaise*, *Finding related to attentiveness*, *Headache*, *Migraine* (*with* and *without aura*), and *Anxiety disorder*. *Neurosis* is also present, but it should be noted that site-labeled source codes for this are almost entirely ICD-10-CM F48.9, *Non-psychotic mental disorder, unspecified* or similar (F48.8 and ICD-9 300.9). Notably, *Impaired cognition* is more common in PASC patients (OR 4.26) but less common in COVID patients (OR 0.53) compared to Controls. Other neurological conditions increased in PASC include *Inflammatory disease of the central nervous system*, *Disorder of autonomic nervous system*, *Polyneuropathy*, *Orthostatic hypotension*, and *Familial dysautonomia* (a genetic condition–see Discussion).

The significant results for PASC also highlight a variety of symptoms related to the cardiovascular, pulmonary, and immune systems. Cardiac conditions such as *Tachycardia*, *Palpitations*, *Congestive heart failure*, *Myocarditis*, *Cardiomyopathy*, and *Cardiomegaly* are observed. Pulmonary issues are well represented with *Pulmonary embolism*, *Bronchiectasis*, *Fibrosis of lung*, and various generic labels for respiratory failure or disorder. Amongst immunological conditions are *Reactive arthritis triad*, *Elevated C-reactive protein*, *Lymphocytopenia*, *Hypogammaglobulinemia*, *Systemic mast cell disease*, and generic *Immunodeficiency disorder*. In addition, bacterial, viral, and fungal infections are increased, including *Bacterial infection due to Pseudomonas*, *Aspergillosis*, and *Pneumocystosis*. Other common themes include musculoskeletal issues (*Fibromyalgia*, *Muscle weakness*, various types of pain) and hematological issues (*Blood coagulation disorder*, *Anemia*, *Hypocalcemia*, *Hypokalemia*).

The analysis also reveals estimated odds ratios less than 1, indicating decreased incidence post-infection compared to Controls, for 219 conditions in one or both cohorts. Most of these (174) were significant only for the larger COVID cohort, and several are related to routine screening or elective procedures potentially disrupted by a COVID-19 infection or lack of care access during the pandemic, such as *Pre-operative state*, *Nicotine dependence*, *Radiological finding*, *Gonarthrosis*, and *Hypertensive disorder*.^43^ *Preoperative state* was largely coded as SNOMED CT 72077002 or ICD-10-CM Z01.818, both widely used across sites and indicative of pre-surgical examination. *Unable to Assess Risk* appears to be a custom code used by a single site, mapped to OMOP concept ID 42690761 by N3C. Other conditions may be more difficult to identify in the six months after a COVID-19 infection due to symptom masking or altered care-seeking behavior. Examples include *Diverticulosis of large intestine* and *Esophageal dysphagia*.^44,45^ In addition to *Pre-operative state*, five conditions are significantly decreased for PASC patients, all related to late-term pregnancy, while *Third trimester pregnancy* is increased in COVID patients (see Discussion).

### Topics significant for PASC and COVID by demographic

From the assessment set, 2,859 PASC patients, 89,374 COVID patients, and 303,017 Control patients met cohort eligibility criteria for per-topic logistic models; Suppl. Table 5 provides per-group patient counts. Baseline contrasts broadly reflected expected trends by life stage and sex (Suppl. Figure 3). T-2 for example pertains to pregnancy, with an estimated female/male OR of 45, pediatric/adult OR 0.06, adolescent/adult 0.2, and senior/adult 0.03. Similarly, T-3 highly weights neonatal conditions and generates a pediatric/adult OR of 43, but no significant female/male trend.

Our primary contrasts considered life stage, sex, and infection-wave demographic groups, evaluating post-vs-pre topic odds radios for PASC or COVID patients compared to corresponding odds ratios for Controls. For example, the contrast ((PASC adult post) / (PASC adult pre)) / ((Control adult post) / (Control adult pre)) results in an OR estimate of 9.89 for T-23, suggesting that post-infection, adult PASC patients increase their odds of generating conditions from this topic nearly 10 times more than Controls do over a similar timeframe. Figure 4 illustrates this result and others for the subset of topics with significant OR estimates >2 for more than one demographic group. All effectiveness and contrast results are listed in Suppl. Table 6 and visualized in Suppl. Figure 4.

**Figure 4:**
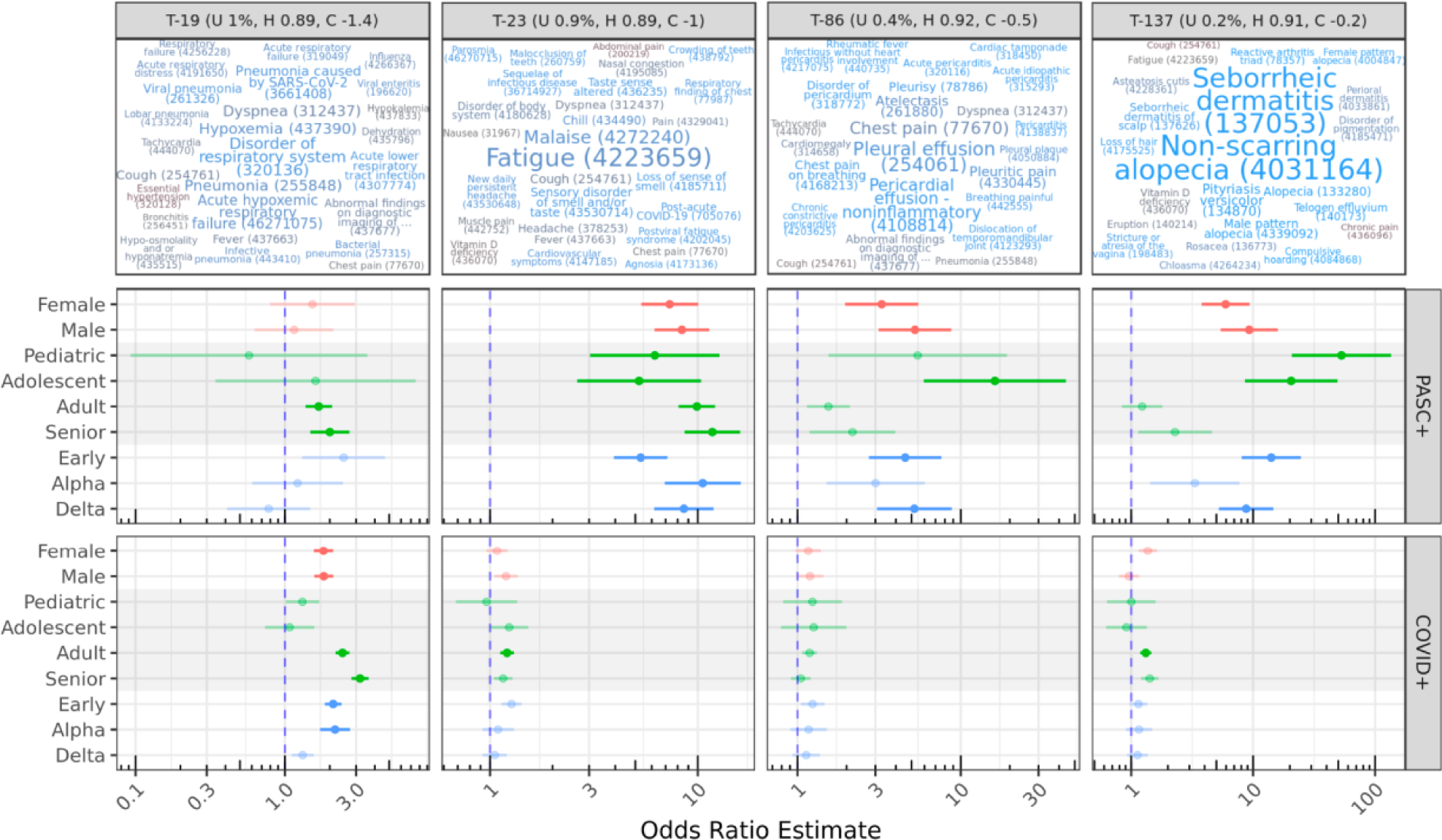
Topics with significant OR estimates >2 for at least two demographic groups. The top row illustrates topics using the same color and size scales as Figure 2; OR estimates are shown for demographic-specific contrasts of PASC or COVID pre-vs-post odds ratios compared to similar Control odds ratios. For example, adult PASC patients increase odds of generating conditions from T-23 post-infection nearly 10 times more than Controls do over a similar timeframe (see Results). Lines show 95% confidence intervals for estimates; semi-transparent estimates are shown for context but were not significant after multiple-test correction.

Amongst the 5,400 sex, life-stage, and wave-specific contrasts, 314 are significant after multiple correction, representing 68 distinct topics. Of these, 130 are represented by the final 10 low quality topics with OR ∼0.6 for all patient groups, potentially reflecting broad healthcare access patterns driven given their shared similarity and few high-relevance terms. Most contrasts have small ORs, with only 30 contrasts across 9 topics having an OR of 2 or higher. The majority of strong effects are seen for the PASC cohort, and while topic coherence was largely uncorrelated with PASC or COVID association, topics with the strongest significant increases in the PASC cohort were less coherent than average (Suppl. Figure 8). PASC confidence intervals were larger due to this cohort’s much smaller size, a trend also seen across relative group sizes.

T-23 stands out as a topic with strong migration among PASC patients, with all subgroups having significant estimated ORs of 5-10. High-weight, high-relevance conditions in T-23 include *Fatigue*, *Malaise*, *Loss of sense of smell*, and other well-known PASC symptoms, as well as the diagnosis code for PASC itself (Post-acute COVID-19). By contrast, COVID patients do not show statistically significant migration to this topic, with the exception of Adults with a small OR of 1.2.

T-19 shows significant OR estimates for several PASC and COVID groups with similar magnitudes. This topic includes several variants of pneumonia and acute respiratory infection symptoms (*Disorder of respiratory system*, *Dyspnea*, *Hypoxemia*, *Cough*), suggesting significant long-term COVID-19 or secondary infections at least 45 days post-primary-infection. For both PASC and COVID cohorts, these increases are most associated with early-wave infections.

Topics 86 and 137 show increases for several PASC groups, especially pediatric and adolescent patients. While T-86 is characterized by *Pleural* and *Pericardial effusion* and related pain, T-137 describes skin conditions, particularly hair loss, including *Non-scarring alopecia* and *Telogen effluvium*, both identified individually above. While effusion is a known factor for severe COVID-19 pneumonia, especially in older patients,^46^ these results highlight similar outcomes in young patients. A systematic review of alopecia in COVID-19 patients by Nguyen and Tosti found that *Anagen effluvium* was associated with younger patients compared to other types of alopecia, but few of the reviewed studies included young patients.^47^

Figure 5 displays additional results for selected topics with cohort or demographic-specific patterns. T-8 represents cardiovascular conditions, and shows a mild but significant increase for adult COVID patients compared to controls. T-43 (not shown) is also significant for PASC adult patients, and encompasses pulmonary conditions. Several of the top-weighted conditions within these topics were individually significant, such as *Palpitations*, *Cardiac arrhythmia*, *Chronic obstructive lung disease*, and *Pulmonary emphysema* for both cohorts, and for PASC *Dizziness and giddiness* and *Tachycardia*. While all of these were individually increased in the PASC cohort, *Cardiac arrhythmia, Chronic obstructive lung disease*, and *Pulmonary emphysema* were decreased in the COVID cohort relative to controls.

**Figure 5:**
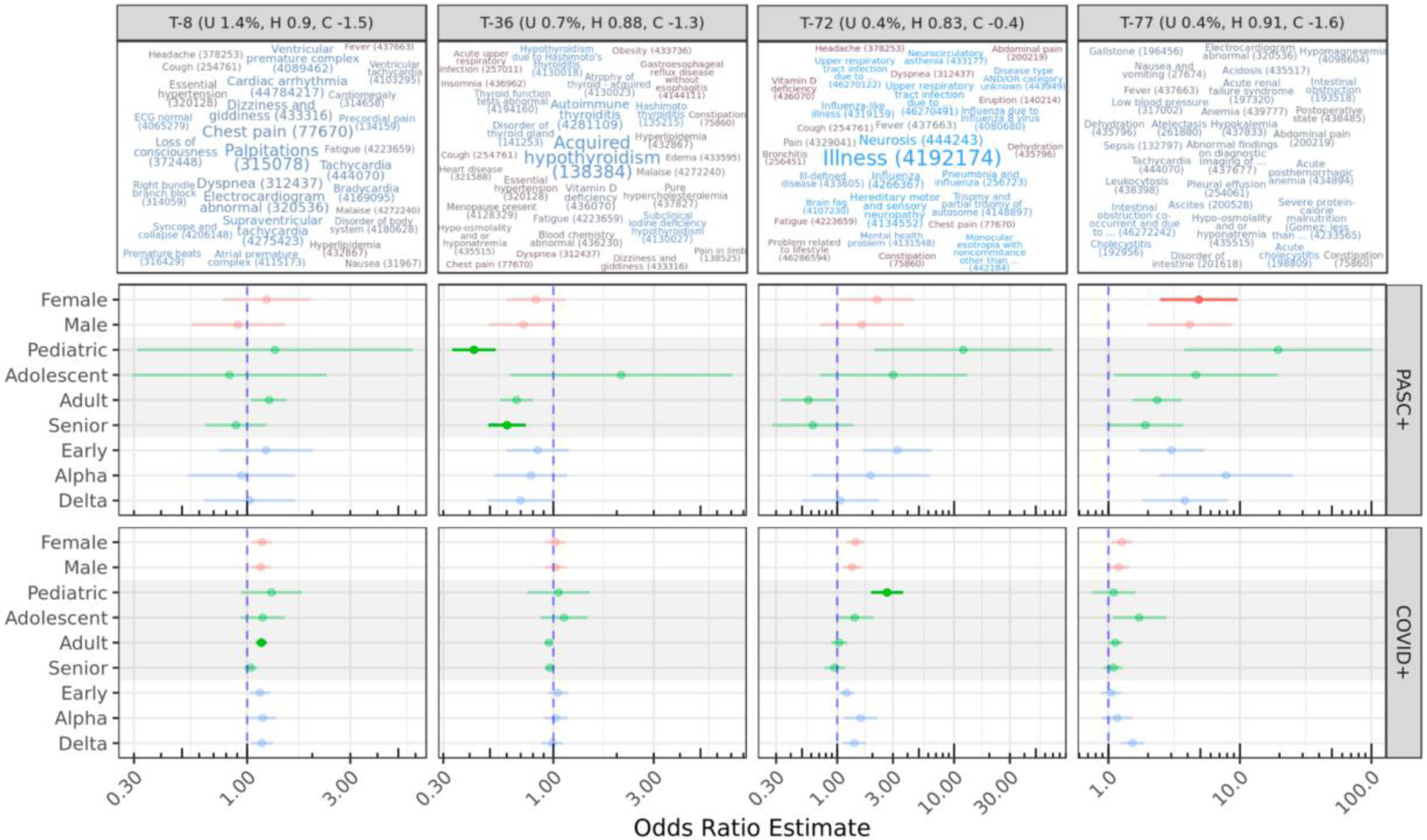
Other select topics with demographic or cohort-specific trends. T-8 is statistically significant only for COVID adults compared to controls. Topics 72 and 77 include diffuse sets of conditions, while T-36 is reduced for PASC pediatric and senior patients, despite representing known PASC outcomes (see Discussion).

T-72 is increased for both COVID and PASC pediatric patients compared to Controls, though this is only statistically significant for the larger COVID cohort. It covers a range of non-specific PASC-like conditions, including *Illness*, *Neurosis* (also discussed above), *Ill-defined disease*, *Mental health problem*, and *Disease type and/or category unknown*. *Brain fog* and *Neurocirculatory asthenia* are additionally found in this topic.

T-77 is increased in female PASC patients compared to controls. This topic is diffuse and has no particularly highly weighted conditions, although many had high relevance scores to the topic. Several of these are laboratory-based, such as *Hypokalemia*, *Anemia*, and *Hyponatremia*. *Tachycardia*, *Pleural effusion*, *Deficiency of macronutrients*, and *Adult failure to thrive syndrome* are also present. The low specificity and coherence of T-77 make it difficult to interpret, although many of these conditions were individually significant above. T-20 (not shown) was increased for COVID adults and COVID delta-wave patients, and also has few high-weight terms, but relevant conditions include *Acute renal failure syndrome*, *Sepsis*, and *Acidosis*.

T-36 strongly decreased for both pediatric and senior PASC patients, and covers only a few conditions with high weights and relevance scores, including *Acquired hypothyroidism* and *Autoimmune thyroiditis*. This result is paradoxical, as these conditions are common long-term outcomes of COVID-19 infection.^48^ Another paradoxical result is a strong (OR 11.7) increase in T-92 for adolescent PASC patients, which covers a variety of physical contusions, lacerations, and abrasions. The highest-weighted condition in this topic however is *Traumatic and/or non-traumatic injury*, all of which were originally coded as ICD-10 T14.8 *Other injury of unspecified body region* for these patients.

Adolescent PASC patients are increased in four topics: T-23, T-86, and T-137 already discussed, and T-174 which highly weights *Thyrotoxicosis*, *C-reactive protein abnormal*, and *Polymyalgia rheumatica*. PASC pediatric patients increase significantly in T-23 and T-137 already discussed, as well as T-57 covering a variety of pulmonary issues such as *Chronic cough*, *Bronchiectasis*, and *Hemoptysis*. On the other hand, PASC adolescent patients were reduced in seven topics and PASC pediatric patients showed a reduction in sixteen, covering a broad range of conditions. These assessment cohorts are small, with 49 pediatric and 66 adolescent patients. Chart reviews revealed that they were distributed across 18 and 20 sites, respectively, and had a similar mean number of conditions recorded in the year prior to infection as other cohorts in the same life stages. However, mean condition counts for these PASC patients were nearly 50% higher in the 6-month post-infection phase (Suppl. Table 5).

These models included covariates to account for site-level differences in topic usage, percentage of PASC patients, and source common data model. To assess the importance of these, we also ran models without them for the subset of topics shown in Figures 4 and 5. Results are highly similar (Suppl. Figure 9), with models without site-level covariates showing slightly higher (< 6%) odds ratios for topics 23, 36, and 72.

## Discussion

While an ICD-10-CM diagnosis code (U09.9) and specialty clinics exist to treat Long COVID, there is still work to be done identifying PASC conditions and how these new diagnoses and referrals are used in practice.^28,49^ Our model, trained on 387 million condition records from 8.9 million patients in the N3C, is one of the most extensive applications of topic modeling to EHR data to date, generating hundreds of diverse and clinically-relevant topics. Only a handful of topics were of low quality, and those in the middle by usage tended to have the highest coherence scores. We hypothesize that common topics are encumbered by a variety of coding options and practices, while rare topics support only a few relevant conditions on top of more common and unrelated background conditions. We found these trends across models with different topic counts, potentially driven by the use of Dirichlet distributions initialized with sparse uniform priors. Topic usage and coherence varied across contributing sites, with notable patterns of usage at PEDsnet sites in particular. Topic modeling may provide insights into site differences in coding practices or data quality, which are concerns in federated and centralized data repositories.^42^

Investigating top-weighted topic terms revealed many conditions associated with increased new-onset rates in PASC and COVID cohorts compared to Controls, including neurocognitive, cardiovascular, pulmonary, and immune-related. Most of these were significant for both cohorts or only the PASC cohort, despite its smaller size. A number of conditions showed lower new incidence in COVID patients compared to Controls, possibly due to decreased access to routine care (e.g. breast cancer^50^) or behavioral changes (e.g. diverticulosis^44^) through the pandemic.

Modeling patient-topic assignment supports queries across patient demographics at a topic level. This approach identified several topics increasing in PASC and COVID patient groups relative to Controls. T-23 stands out as the clearest PASC-related topic across demographics, and includes many conditions commonly associated with Long COVID such as fatigue, malaise, new daily headache, and dyspnea. Other topics are demographic-specific, such as T-86 covering *Pleural* and *Pericardial effusion*, T-137 with *Non-scarring alopecia* and *Seborrheic dermatitis*, and T-57 covering other pulmonary issues for younger PASC patients.

While most effects are larger for PASC patients, T-19 shows similar effect sizes for COVID adults and seniors. This topic largely represents secondary pneumonias and related symptoms, suggesting that while these are not used as indicators for PASC, they are nevertheless long-term issues for COVID-19 patients. The association is strongest with the early waves of the pandemic, reflecting severity of illness and lack of effective treatment protocols during this period.^51^ Few such wave effects were significant overall; T-20 with *Acute renal failure syndrome*, *Acidosis*, and *Sepsis* is an exception showing increases for COVID delta-wave patients. Despite the few PASC pediatric patients and wide confidence interval ranges, several topics were increased for this group indicating a unique cohort with significant long-term COVID-19 health outcomes. On the other hand, estimates for COVID-only pediatric patients for most topics, including T-23, T-57, and T-137, are non-significant despite a larger sample size.

While this study reaffirms many known PASC trends, several results merit further investigation. Female PASC patients increased in T-77, which is diffuse, multisystem, and covers many conditions identified in other tests. More targeted analyses of this set may reveal a unique sub-phenotype or mix of sub-phenotypes experienced by a unique population. Additionally, T-72 represents a cluster of ill-defined conditions; its increase for COVID pediatric patients may reflect difficulties in PASC identification for this group. For example, the highest-weighted term, *Illness*, was originally coded as ICD-10 R69 *Illness, unspecified* in the vast majority of cases. Amongst individual conditions, *Impaired cognition* increased in PASC patients but decreased in COVID patients. Many of these were originally coded as R41.844, *Frontal lobe and executive function disorder*. Executive dysfunction has been linked to COVID-19, particularly for patients with acute respiratory distress syndrome.^52^ This diagnosis is distinct from those typical for ADHD (F90), so it is unclear whether the reduction observed in COVID patients was a result of reduced healthcare access.

In contrast to other studies,^53,54^ we found few gastrointestinal conditions increased in PASC or COVID patients, though *Abdominal pain, Viral gastroenteritis*, and *Dysphagia* were increased in PASC patients (Suppl. Table 4). Neither did we find statistically significant sex differences, despite a known increased risk for PASC in female patients. Our experiments, however, evaluate cohorts defined by PASC diagnosis. While female patients are more likely to develop PASC, our results suggest minimal sex differences amongst patients who have been positively identified. Still, other work has suggested sex differences,^55^ and similar non-significant trends in our results may be worthy of followup. The apparent reduction in late-term pregnancy conditions for PASC patients and simultaneous increase for COVID patients (both in comparison to Controls) is notable. We hypothesize that pregnant patients are less likely to be diagnosed with PASC given the similarity of presentation, but more likely to be monitored if infected during pregnancy.

A high incidence of postural orthostatic tachycardia syndrome (POTS) has been identified in PASC clinical research,^56^ but a POTS-specific ICD-10 code did not exist prior to October 1, 2022, and therefore POTS is not present in our dataset. The closest available term in the SNOMED hierarchy, *Orthostatic hypotension*, was found to be significantly elevated in PASC, as were *Disorder of the autonomic nervous system* and *Familial dysautonomia*. Many symptoms significant for the PASC cohort, such as *Tachycardia*, *Palpitations*, *Dizziness and giddiness*, *Fatigue*, and *Finding related to attentiveness* are suggestive of POTS or similar forms of dysautonomia. The presence of *Familial dysautonomia* (ICD-10-CM G90.1), a rare genetic disorder, is unlikely to be due to increased screening given that we saw no corresponding uptake in genetic testing. Rather, we suspect that frequent mis-coding may occur because the ICD-10-CM catalog has only one match for the term “dysautonomia” (G90.1 *Familial dysautonomia*), which when used alone encompasses multiple PASC-related conditions.^57^ Such errors are not uncommon when using medical record software.^58^

Many of our results are immune-related, including conditions (*Lymphocytopenia*, *Hypogammaglobulinemia*, *Systemic mast cell disease*) and infections more common in immunocompromised patients (*Aspergillosis*, *Pneumocystosis*). Topic 36 highly weighting *Hypothyroidism* and *Thyroiditis* shows reductions for PASC pediatric and senior patients, a paradoxical result given that these are known post-acute sequelae.^48^ It may be that patients with pre-existing thyroid disorders are underdiagnosed for PASC, while new thyroid disorders after COVID-19 infection are identified as PASC and related symptoms alone. Together these results suggest an important role for thyroid-mediated dysfunction in PASC patients, and we recommend investigation into how these related diseases are diagnosed and treated.

Design choices and limitations of this study should be considered when interpreting results. To maximize the number and specificity of testable topics, we trained our model on a diverse set of patients with and without COVID-19 and PASC, using complete patient histories to maximize effective document size. LDA does not model temporal relationships between terms when generating topics, and topics may thus highly weigh both risk factors and outcomes. Like many clustering methods, LDA and its online variant are subject to suboptimal convergence resulting in possible variation in topics across runs.^59,60^ To mitigate these risks we employed hyperparameter tuning, increased training iterations, and model evaluation via coherence on an independent validation set. In addition to data filtering for quality and fitness of use, we removed diagnoses for COVID-19 itself prior to topic modeling. Because these conditions largely define inclusion criteria for both N3C and our COVID cohort, they are broadly correlated and their inclusion would likely influence topic composition significantly.

While many results are shared between the COVID and PASC cohorts, it’s important to note that these are computed against a common Control cohort rather than between PASC and COVID directly, and the larger size of the COVID cohort results in increased sensitivity. Overall trends in healthcare utilization and access during the pandemic should be considered, and these may be influenced by COVID-19 infection itself. N3C’s observational EHR data, although extensive, are not a random sample and represent a diversity of specialties, coding practices, and other factors, as evidenced by variation in topic usage and coherence across sites. We excluded patients from sites without any U09.9 PASC diagnoses to minimize misclassification of PASC patients, but these sites may serve unique populations or use topics in distinct ways. Topic-level models included several site-level covariates, including per-site topic usage and percentage of PASC patients. Models excluding these covariates yielded highly similar results, supporting cross-site generalizability, and the use of a held-out assessment set for these tests provides a level of independence from topic generation. Finally, we’ve focused on group-level inferential analyses to broadly understand PASC sub-phenotypes. Although topic models provide per-patient topic associations, the probabilistic nature of LDA limits its utility for individual patients and further research is needed prior to patient-level predictive applications.

With this context in mind, topic modeling applied to a large EHR dataset has proven highly effective for assessing the progression of post-acute sequelae of SARS-CoV-2 infection. Our LDA model identified hundreds of fine-grain potential sub-phenotypes in the data, and interpreting the probabilistic assignment of patients to them through GEE-based logistic regression is a novel and flexible approach, supported by properties of both methods and empirically by expected demographic baselines. Future investigations may assess other factors such as acute disease severity, contrast different cohorts, analyze inter-topic patterns to uncover sub-phenotype-specific risk factors, or employ time-series techniques to examine topic distributions across multiple time windows.

Ultimately, a finer understanding of presentations across populations can inform research, diagnostics, treatment, and health equity for multi-faceted diseases such as PASC. Tracking patient clinical trajectories over time in light of model-derived sub-phenotypes revealed post-acute sequelae of SARS-CoV-2 infection, several of which were associated with patient sex, age, wave of infection, or presence of a PASC diagnosis. Some results, such as those highlighting immune dysfunction, thyroid involvement, and secondary infections improve our understanding of potential mechanisms for PASC. Others, such as those highlighting non-specific phenotypes in the COVID cohort, may lead to improved diagnostics and support for patients suffering from Long-COVID but yet to receive a PASC diagnosis.

## Author Contributions

Concept and modeling: S.O., C.M. Statistical design: S.O., K.W., B.M., J.L. Results interpretation: S.O., H.D., G.A., H.W., M.L., M.H. Data curation: S.O., P.Z., E.F., J.L., A.Z., R.M., E.P., Y.Y., P.L., R.C. Manuscript drafting: S.O., C.M., B.M., J.M., M.H. Material and administrative support: J.M., M.H.

## Data Availability

The N3C Data Enclave is managed under the authority of the NIH; information can be found at https://ncats.nih.gov/n3c/resources. The N3C data transfer to NCATS is performed under a Johns Hopkins University Reliance Protocol # IRB00249128 or individual site agreements with NIH. Enclave data is protected, and can be accessed for COVID-related research with an approved (1) IRB protocol and (2) Data Use Request (DUR). Enclave and data access instructions can be found at https://covid.cd2h.org/for-researchers.

## Code Availability

Analysis code is available at https://github.com/oneilsh/lda_pasc.

## Acknowledgements

We sincerely thank the reviewers for their insightful comments, which significantly enhanced the generalizability and readability of this manuscript.

The analyses described in this publication were conducted with data and tools accessed through the NCATS N3C Data Enclave https://covid.cd2h.org and N3C Attribution & Publication Policy v 1.2-2020-08-25b supported by NCATS U24 TR002306, Axle Informatics Subcontract: NCATS-P00438-B, National Institutes of Health grant NHLBI RECOVER Agreement OT2HL161847-01, and CTSA award No. UM1TR004360 from the National Center for Advancing Translational Sciences. This research was possible because of the patients whose information is included within the data and the organizations (https://ncats.nih.gov/n3c/resources/data-contribution/data-transfer-agreement-signatories) and scientists who have contributed to the on-going development of this community resource [https://doi.org/10.1093/jamia/ocaa196]. This study is part of the NIH Researching COVID to Enhance Recovery (RECOVER) Initiative (https://recovercovid.org/), which seeks to understand, treat, and prevent the post-acute sequelae of SARS-CoV-2 infection (PASC), and was conducted under the N3C DUR RP-5677B5. We would like to thank the National Community Engagement Group (NCEG), all patient, caregiver and community Representatives, and all the participants enrolled in the RECOVER Initiative. Authorship was determined using ICMJE recommendations.

The N3C Publication committee confirmed that this manuscript is in accordance with N3C data use and attribution policies; however, this content is solely the responsibility of the authors and does not necessarily represent the official views of the RECOVER or N3C programs or the National Institutes of Health.

Use of the N3C data for this study is authorized under the following IRB Protocols:

| Site | IRB Name | Exempted vs Approved | Protocol Number |
| --- | --- | --- | --- |
| University of Colorado | Colorado Multiple Institutional Review Board | approved | 21-2759 |
| Johns Hopkins University | Johns Hopkins Office of Human Subjects Research - Institutional Review Board | approved | IRB00249128 |
| University of North Carolina | University of North Carolina Chapel Hill Institutional Review Board | exempt | 21-0309 |
| Stony Brook University | Office of Research Compliance, Division of Human Subject Protections, Stony Brook University | exempt | IRB2021-00098 |
| OCHIN | Advarra IRB | approved | Pro00060719 |
| RTI International | RTI Office of Research Protection | exempt | MOD10001700 |

We gratefully acknowledge the following core contributors to N3C:

Adam B. Wilcox, Adam M. Lee, Alexis Graves, Alfred (Jerrod) Anzalone, Amin Manna, Amit Saha, Amy Olex, Andrea Zhou, Andrew E. Williams, Andrew Southerland, Andrew T. Girvin, Anita Walden, Anjali A. Sharathkumar, Benjamin Amor, Benjamin Bates, Brian Hendricks, Brijesh Patel, Caleb Alexander, Carolyn Bramante, Cavin Ward-Caviness, Charisse Madlock-Brown, Christine Suver, Christopher Chute, Christopher Dillon, Chunlei Wu, Clare Schmitt, Cliff Takemoto, Dan Housman, Davera Gabriel, David A. Eichmann, Diego Mazzotti, Don Brown, Eilis Boudreau, Elaine Hill, Elizabeth Zampino, Emily Carlson Marti, Emily R. Pfaff, Evan French, Farrukh M Koraishy, Federico Mariona, Fred Prior, George Sokos, Greg Martin, Harold Lehmann, Heidi Spratt, Hemalkumar Mehta, Hongfang Liu, Hythem Sidky, J.W. Awori Hayanga, Jami Pincavitch, Jaylyn Clark, Jeremy Richard Harper, Jessica Islam, Jin Ge, Joel Gagnier, Joel H. Saltz, Joel Saltz, Johanna Loomba, John Buse, Jomol Mathew, Joni L. Rutter, Julie A. McMurry, Justin Guinney, Justin Starren, Karen Crowley, Katie Rebecca Bradwell, Kellie M. Walters, Ken Wilkins, Kenneth R. Gersing, Kenrick Dwain Cato, Kimberly Murray, Kristin Kostka, Lavance Northington, Lee Allan Pyles, Leonie Misquitta, Lesley Cottrell, Lili Portilla, Mariam Deacy, Mark M. Bissell, Marshall Clark, Mary Emmett, Mary Morrison Saltz, Matvey B. Palchuk, Melissa A. Haendel, Meredith Adams, Meredith Temple-O’Connor, Michael G. Kurilla, Michele Morris, Nabeel Qureshi, Nasia Safdar, Nicole Garbarini, Noha Sharafeldin, Ofer Sadan, Patricia A. Francis, Penny Wung Burgoon, Peter Robinson, Philip R.O. Payne, Rafael Fuentes, Randeep Jawa, Rebecca Erwin-Cohen, Rena Patel, Richard A. Moffitt, Richard L. Zhu, Rishi Kamaleswaran, Robert Hurley, Robert T. Miller, Saiju Pyarajan, Sam G. Michael, Samuel Bozzette, Sandeep Mallipattu, Satyanarayana Vedula, Scott Chapman, Shawn T. O’Neil, Soko Setoguchi, Stephanie S. Hong, Steve Johnson, Tellen D. Bennett, Tiffany Callahan, Umit Topaloglu, Usman Sheikh, Valery Gordon, Vignesh Subbian, Warren A. Kibbe, Wenndy Hernandez, Will Beasley, Will Cooper, William Hillegass, Xiaohan Tanner Zhang. Details of contributions available at covid.cd2h.org/core-contributors

The following institutions whose data is released or pending:

Available: Advocate Health Care Network — UL1TR002389: The Institute for Translational Medicine (ITM) • Aurora Health Care Inc — UL1TR002373: Wisconsin Network For Health Research • Boston University Medical Campus — UL1TR001430: Boston University Clinical and Translational Science Institute • Brown University — U54GM115677: Advance Clinical Translational Research (Advance-CTR) • Carilion Clinic — UL1TR003015: iTHRIV Integrated Translational health Research Institute of Virginia • Case Western Reserve University — UL1TR002548: The Clinical & Translational Science Collaborative of Cleveland (CTSC) • Charleston Area Medical Center — U54GM104942: West Virginia Clinical and Translational Science Institute (WVCTSI) • Children’s Hospital Colorado — UL1TR002535: Colorado Clinical and Translational Sciences Institute • Columbia University Irving Medical Center — UL1TR001873: Irving Institute for Clinical and Translational Research • Dartmouth College — None (Voluntary) Duke University — UL1TR002553: Duke Clinical and Translational Science Institute • George Washington Children’s Research Institute — UL1TR001876: Clinical and Translational Science Institute at Children’s National (CTSA-CN) • George Washington University — UL1TR001876: Clinical and Translational Science Institute at Children’s National (CTSA-CN) • Harvard Medical School — UL1TR002541: Harvard Catalyst • Indiana University School of Medicine — UL1TR002529: Indiana Clinical and Translational Science Institute • Johns Hopkins University — UL1TR003098: Johns Hopkins Institute for Clinical and Translational Research • Louisiana Public Health Institute — None (Voluntary) • Loyola Medicine — Loyola University Medical Center • Loyola University Medical Center — UL1TR002389: The Institute for Translational Medicine (ITM) • Maine Medical Center — U54GM115516: Northern New England Clinical & Translational Research (NNE-CTR) Network • Mary Hitchcock Memorial Hospital & Dartmouth Hitchcock Clinic — None (Voluntary) • Massachusetts General Brigham — UL1TR002541: Harvard Catalyst • Mayo Clinic Rochester — UL1TR002377: Mayo Clinic Center for Clinical and Translational Science (CCaTS) • Medical University of South Carolina — UL1TR001450: South Carolina Clinical & Translational Research Institute (SCTR) • MITRE Corporation — None (Voluntary) • Montefiore Medical Center — UL1TR002556: Institute for Clinical and Translational Research at Einstein and Montefiore • Nemours — U54GM104941: Delaware CTR ACCEL Program • NorthShore University HealthSystem — UL1TR002389: The Institute for Translational Medicine (ITM) • Northwestern University at Chicago — UL1TR001422: Northwestern University Clinical and Translational Science Institute (NUCATS) • OCHIN — INV-018455: Bill and Melinda Gates Foundation grant to Sage Bionetworks • Oregon Health & Science University — UL1TR002369: Oregon Clinical and Translational Research Institute • Penn State Health Milton S. Hershey Medical Center — UL1TR002014: Penn State Clinical and Translational Science Institute • Rush University Medical Center — UL1TR002389: The Institute for Translational Medicine (ITM) • Rutgers, The State University of New Jersey — UL1TR003017: New Jersey Alliance for Clinical and Translational Science • Stony Brook University — U24TR002306 • The Alliance at the University of Puerto Rico, Medical Sciences Campus — U54GM133807: Hispanic Alliance for Clinical and Translational Research (The Alliance) • The Ohio State University — UL1TR002733: Center for Clinical and Translational Science • The State University of New York at Buffalo — UL1TR001412: Clinical and Translational Science Institute • The University of Chicago — UL1TR002389: The Institute for Translational Medicine (ITM) • The University of Iowa — UL1TR002537: Institute for Clinical and Translational Science • The University of Miami Leonard M. Miller School of Medicine — UL1TR002736: University of Miami Clinical and Translational Science Institute • The University of Michigan at Ann Arbor — UL1TR002240: Michigan Institute for Clinical and Health Research • The University of Texas Health Science Center at Houston — UL1TR003167: Center for Clinical and Translational Sciences (CCTS) • The University of Texas Medical Branch at Galveston — UL1TR001439: The Institute for Translational Sciences • The University of Utah — UL1TR002538: Uhealth Center for Clinical and Translational Science • Tufts Medical Center — UL1TR002544: Tufts Clinical and Translational Science Institute • Tulane University — UL1TR003096: Center for Clinical and Translational Science • The Queens Medical Center — None (Voluntary) • University Medical Center New Orleans — U54GM104940: Louisiana Clinical and Translational Science (LA CaTS) Center • University of Alabama at Birmingham — UL1TR003096: Center for Clinical and Translational Science • University of Arkansas for Medical Sciences — UL1TR003107: UAMS Translational Research Institute • University of Cincinnati — UL1TR001425: Center for Clinical and Translational Science and Training • University of Colorado Denver, Anschutz Medical Campus — UL1TR002535: Colorado Clinical and Translational Sciences Institute • University of Illinois at Chicago — UL1TR002003: UIC Center for Clinical and Translational Science • University of Kansas Medical Center — UL1TR002366: Frontiers: University of Kansas Clinical and Translational Science Institute • University of Kentucky — UL1TR001998: UK Center for Clinical and Translational Science • University of Massachusetts Medical School Worcester — UL1TR001453: The UMass Center for Clinical and Translational Science (UMCCTS) • University Medical Center of Southern Nevada — None (voluntary) • University of Minnesota — UL1TR002494: Clinical and Translational Science Institute • University of Mississippi Medical Center — U54GM115428: Mississippi Center for Clinical and Translational Research (CCTR) • University of Nebraska Medical Center — U54GM115458: Great Plains IDeA-Clinical & Translational Research • University of North Carolina at Chapel Hill — UL1TR002489: North Carolina Translational and Clinical Science Institute • University of Oklahoma Health Sciences Center — U54GM104938: Oklahoma Clinical and Translational Science Institute (OCTSI) • University of Pittsburgh — UL1TR001857: The Clinical and Translational Science Institute (CTSI) • University of Pennsylvania — UL1TR001878: Institute for Translational Medicine and Therapeutics • University of Rochester — UL1TR002001: UR Clinical & Translational Science Institute • University of Southern California — UL1TR001855: The Southern California Clinical and Translational Science Institute (SC CTSI) • University of Vermont — U54GM115516: Northern New England Clinical & Translational Research (NNE-CTR) Network • University of Virginia — UL1TR003015: iTHRIV Integrated Translational health Research Institute of Virginia • University of Washington — UL1TR002319: Institute of Translational Health Sciences • University of Wisconsin-Madison — UL1TR002373: UW Institute for Clinical and Translational Research • Vanderbilt University Medical Center — UL1TR002243: Vanderbilt Institute for Clinical and Translational Research • Virginia Commonwealth University — UL1TR002649: C. Kenneth and Dianne Wright Center for Clinical and Translational Research • Wake Forest University Health Sciences — UL1TR001420: Wake Forest Clinical and Translational Science Institute • Washington University in St. Louis — UL1TR002345: Institute of Clinical and Translational Sciences • Weill Medical College of Cornell University — UL1TR002384: Weill Cornell Medicine Clinical and Translational Science Center • West Virginia University — U54GM104942: West Virginia Clinical and Translational Science Institute (WVCTSI) Submitted: Icahn School of Medicine at Mount Sinai — UL1TR001433: ConduITS Institute for Translational Sciences • The University of Texas Health Science Center at Tyler — UL1TR003167: Center for Clinical and Translational Sciences (CCTS) • University of California, Davis — UL1TR001860: UCDavis Health Clinical and Translational Science Center • University of California, Irvine — UL1TR001414: The UC Irvine Institute for Clinical and Translational Science (ICTS) • University of California, Los Angeles — UL1TR001881: UCLA Clinical Translational Science Institute • University of California, San Diego — UL1TR001442: Altman Clinical and Translational Research Institute • University of California, San Francisco — UL1TR001872: UCSF Clinical and Translational Science Institute  Pending: Arkansas Children’s Hospital — UL1TR003107: UAMS Translational Research Institute • Baylor College of Medicine — None (Voluntary) • Children’s Hospital of Philadelphia — UL1TR001878: Institute for Translational Medicine and Therapeutics • Cincinnati Children’s Hospital Medical Center — UL1TR001425: Center for Clinical and Translational Science and Training • Emory University — UL1TR002378: Georgia Clinical and Translational Science Alliance • HonorHealth — None (Voluntary) • Loyola University Chicago — UL1TR002389: The Institute for Translational Medicine (ITM) • Medical College of Wisconsin — UL1TR001436: Clinical and Translational Science Institute of Southeast Wisconsin • MedStar Health Research Institute — None (Voluntary) • Georgetown University — UL1TR001409: The Georgetown-Howard Universities Center for Clinical and Translational Science (GHUCCTS) • MetroHealth — None (Voluntary) • Montana State University — U54GM115371: American Indian/Alaska Native CTR • NYU Langone Medical Center — UL1TR001445: Langone Health’s Clinical and Translational Science Institute • Ochsner Medical Center — U54GM104940: Louisiana Clinical and Translational Science (LA CaTS) Center • Regenstrief Institute — UL1TR002529: Indiana Clinical and Translational Science Institute • Sanford Research — None (Voluntary) • Stanford University — UL1TR003142: Spectrum: The Stanford Center for Clinical and Translational Research and Education • The Rockefeller University — UL1TR001866: Center for Clinical and Translational Science • The Scripps Research Institute — UL1TR002550: Scripps Research Translational Institute • University of Florida — UL1TR001427: UF Clinical and Translational Science Institute • University of New Mexico Health Sciences Center — UL1TR001449: University of New Mexico Clinical and Translational Science Center • University of Texas Health Science Center at San Antonio — UL1TR002645: Institute for Integration of Medicine and Science • Yale New Haven Hospital — UL1TR001863: Yale Center for Clinical Investigation

## Supplemental Figures

**Suppl. Figure 1.**
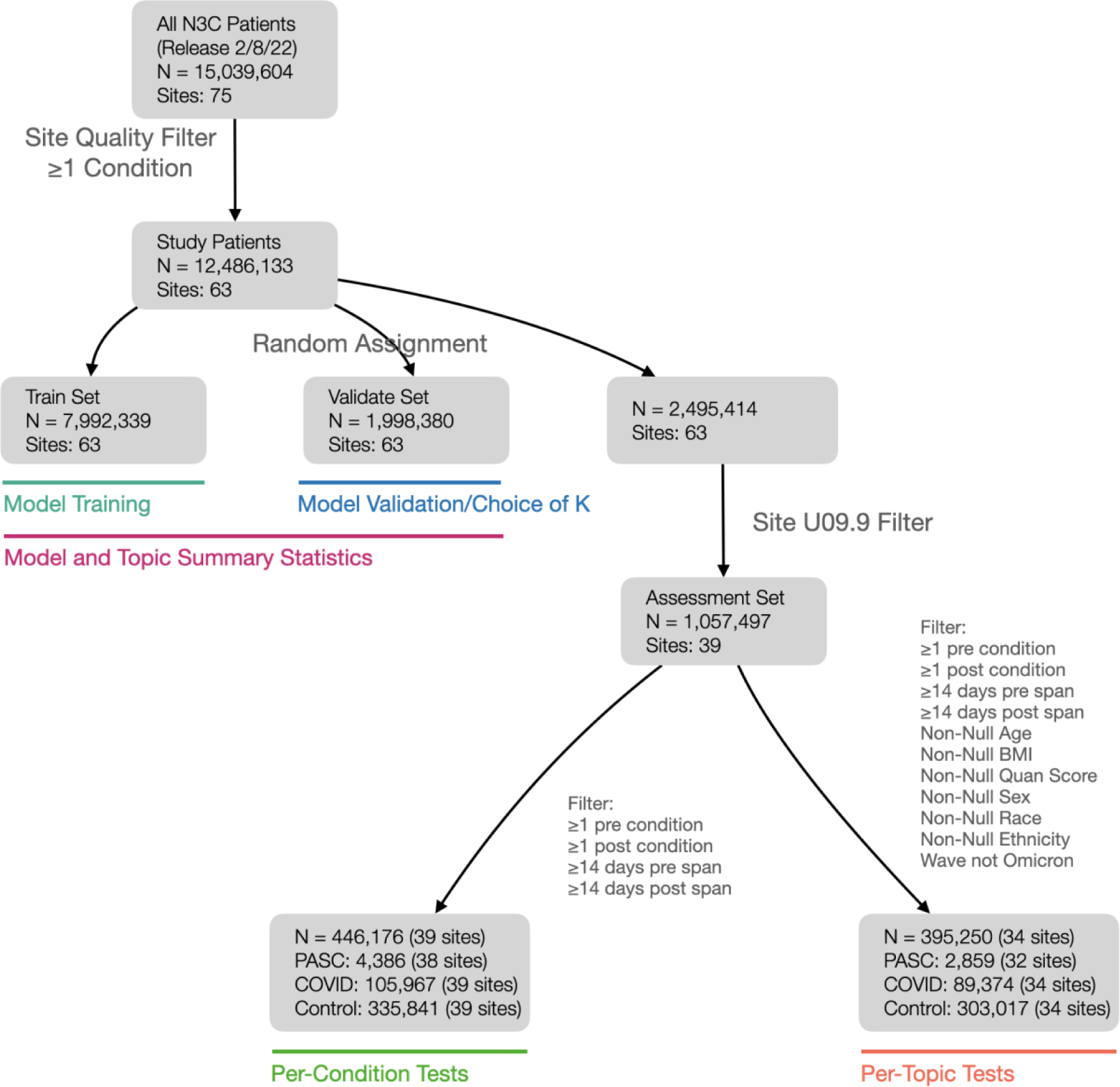
Consort diagram illustrating stratification of patients into sets and cohorts, number of unique sites represented by those groups, and how each is used in analysis. The site quality filter removed sites with inpatient serum creatinine or white blood cell count results for fewer than 25% of patients, the site U09.9 filter removed patients from sites with no U09.9 diagnoses, and filter variables are as described for specific tests (see Suppl. Methods).

**Suppl. Figure 2.**
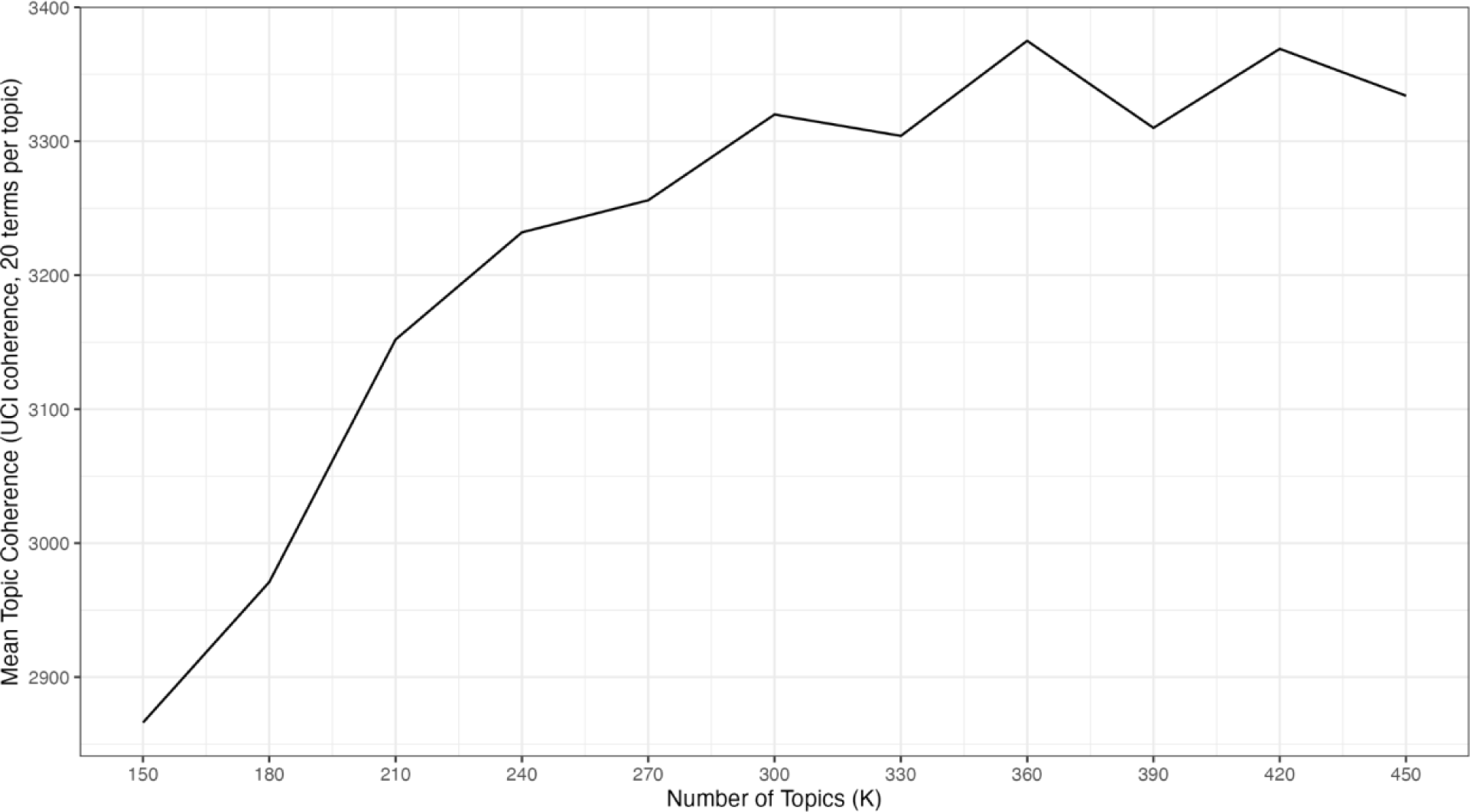
Mean topic coherence scores for LDA models varying the number of topics generated (K). Topic coherences are computed as intrinsic UCI Coherence^30^ using the top 20 terms per topic. UCI coherence evaluates, for all term pairs amongst these top 20, how frequently they occur together in patient histories compared to the expectation assuming terms occur independently, on the validation data set. K=300 was chosen as the final number of topics.

Suppl. Figure 3

Full topic clouds for all 300 topics generated and visualizations of corresponding contrasts. Available at https://doi.org/10.5281/zenodo.11188766.

**Suppl. Figure 4.**
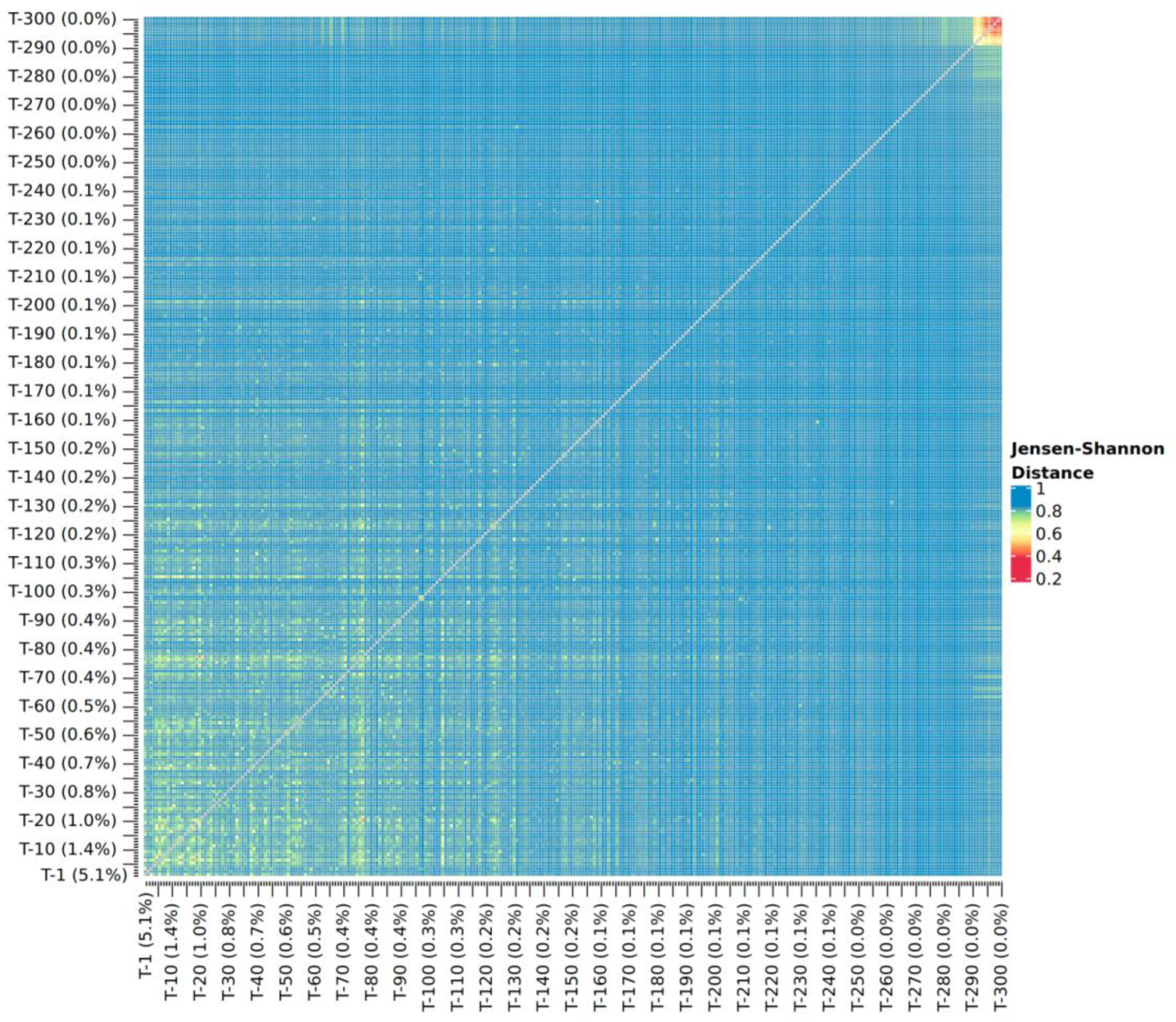
Topic/topic dissimilarity as Jensen-Shannon Distance. Topic self-distances of 0 are not shown.

**Suppl. Figure 5.**
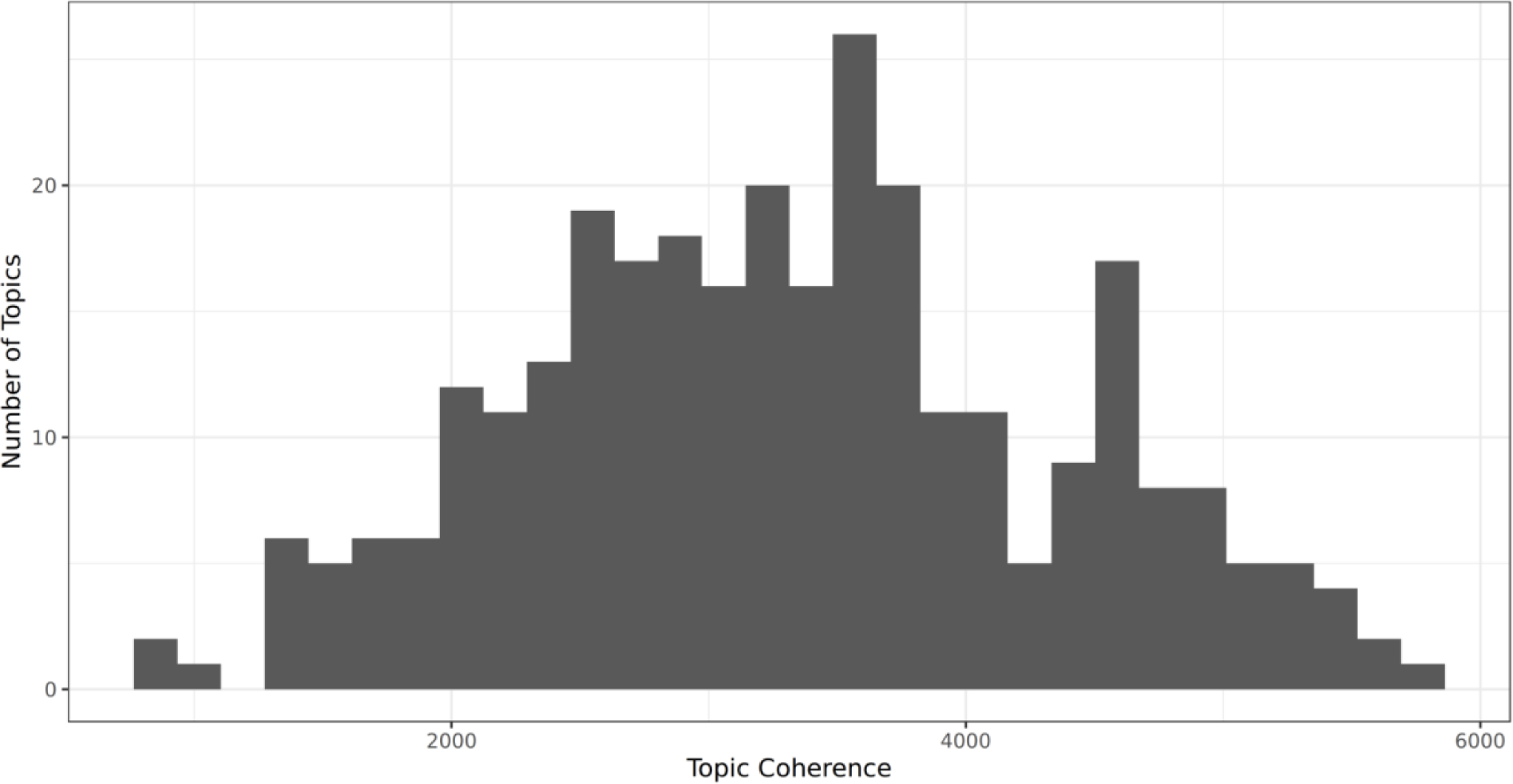
Histogram of topic coherence values.

**Suppl. Figure 6.**
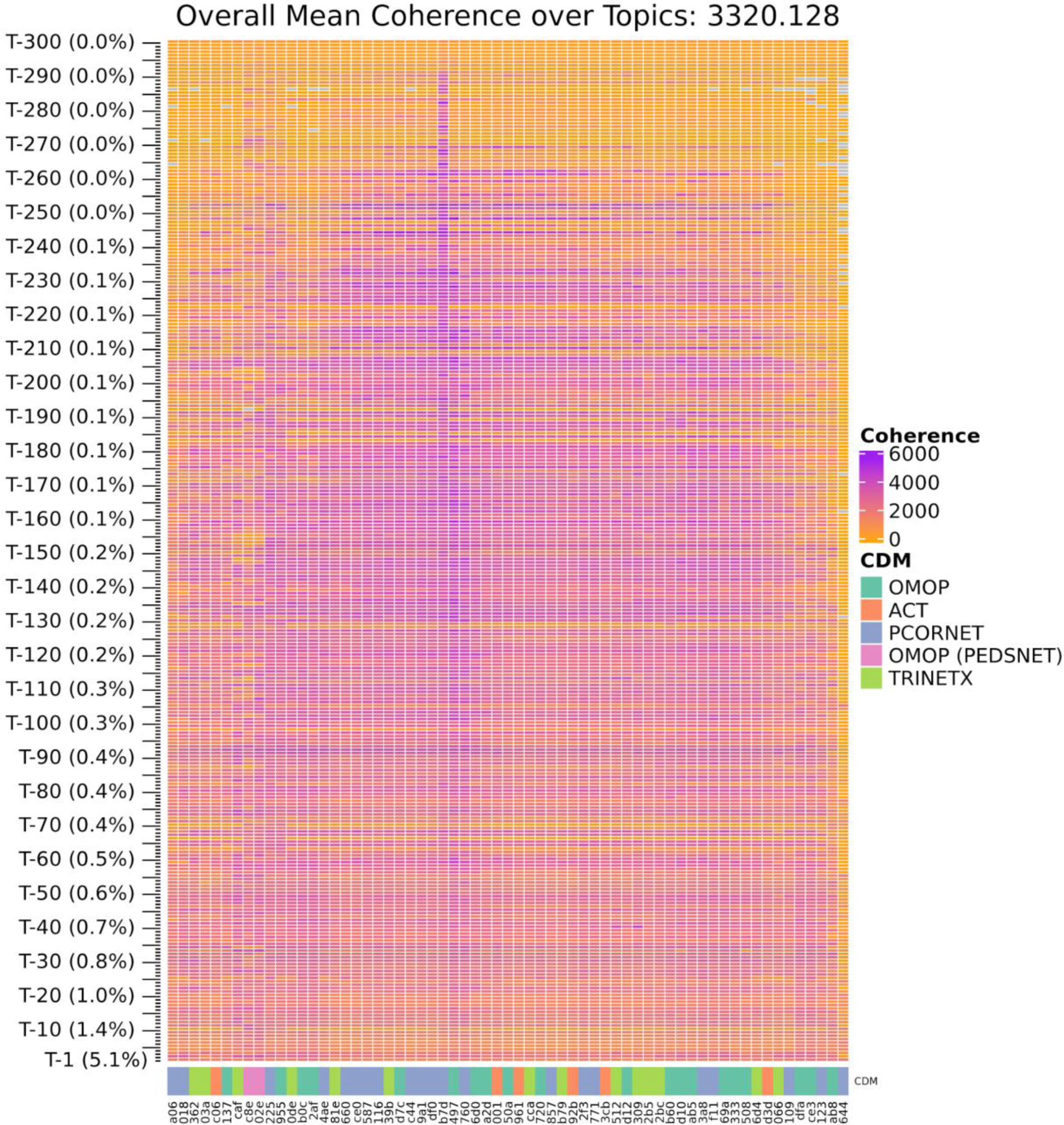
Mean UCI coherence scores per topic and contributing data site (ID anonymized). Site identifiers are masked, but labeled with the source common data model in use at the site.

**Suppl. Figure 7.**
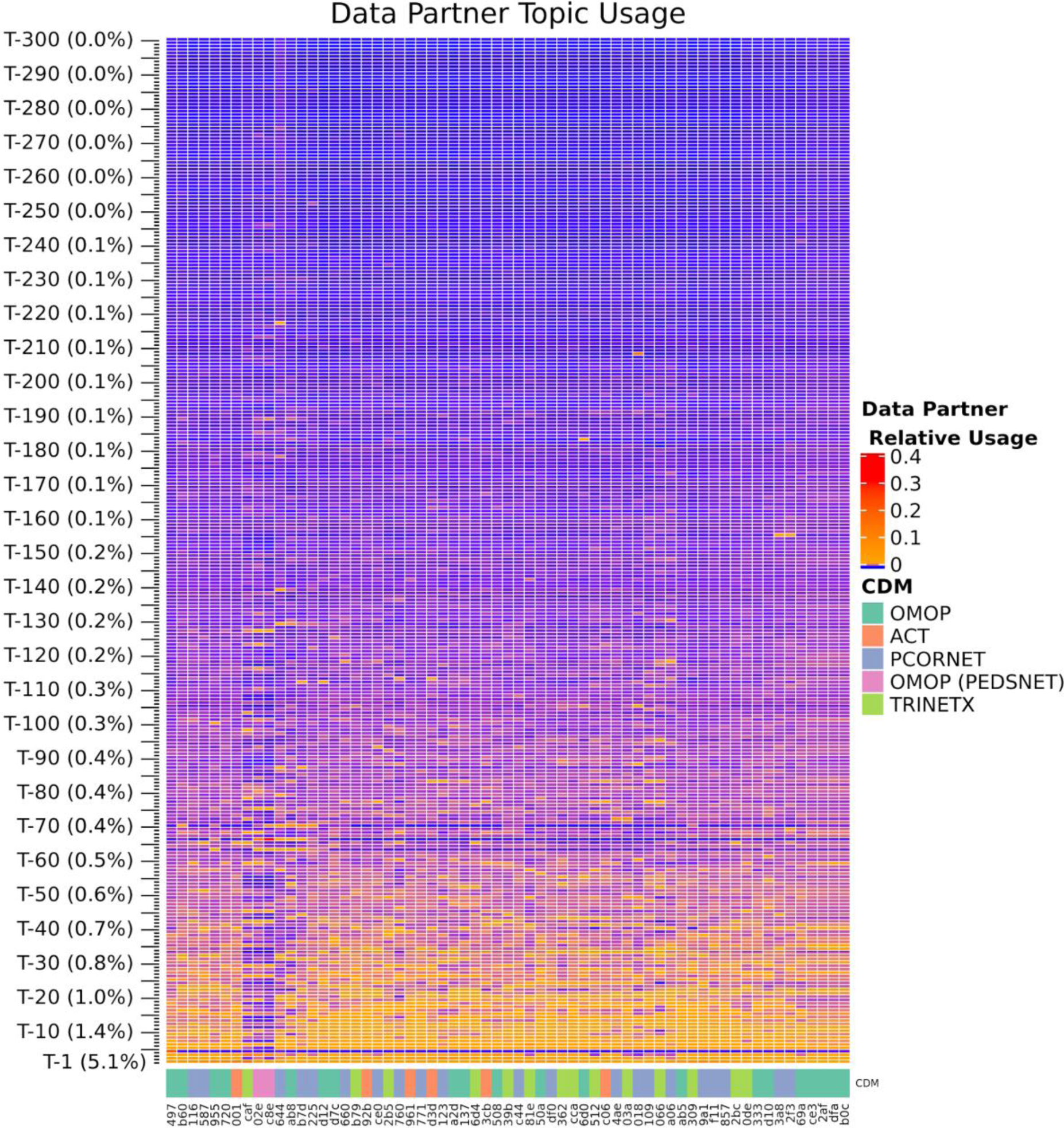
Relative usage of topics per contributing site (ID anonymized). For a given site and topic, relative usage is computed as the sum of assigned weights to that topic for patients from that site divided by the number of patients, representing a distribution over topics per site.

**Suppl. Figure 8.**
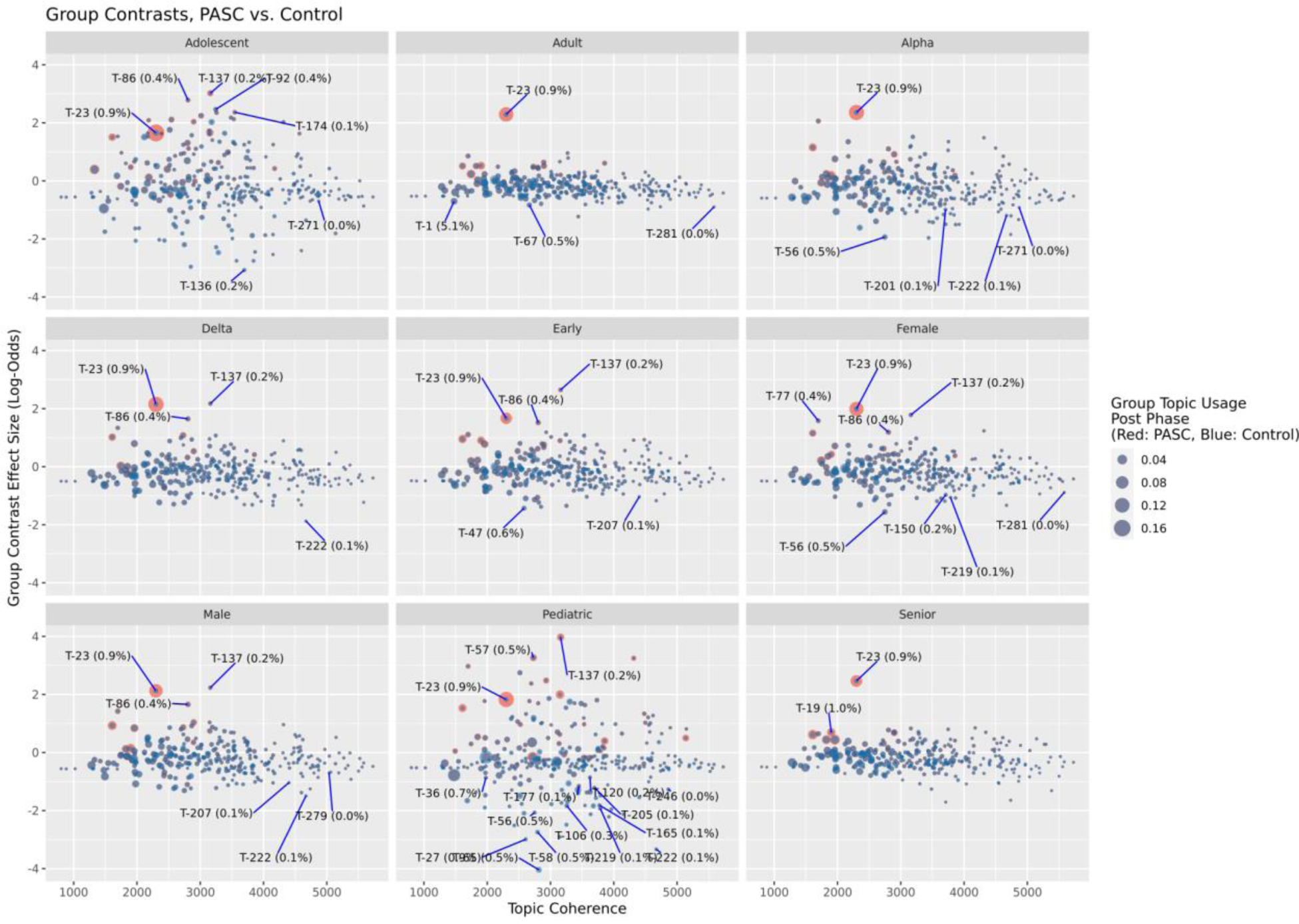
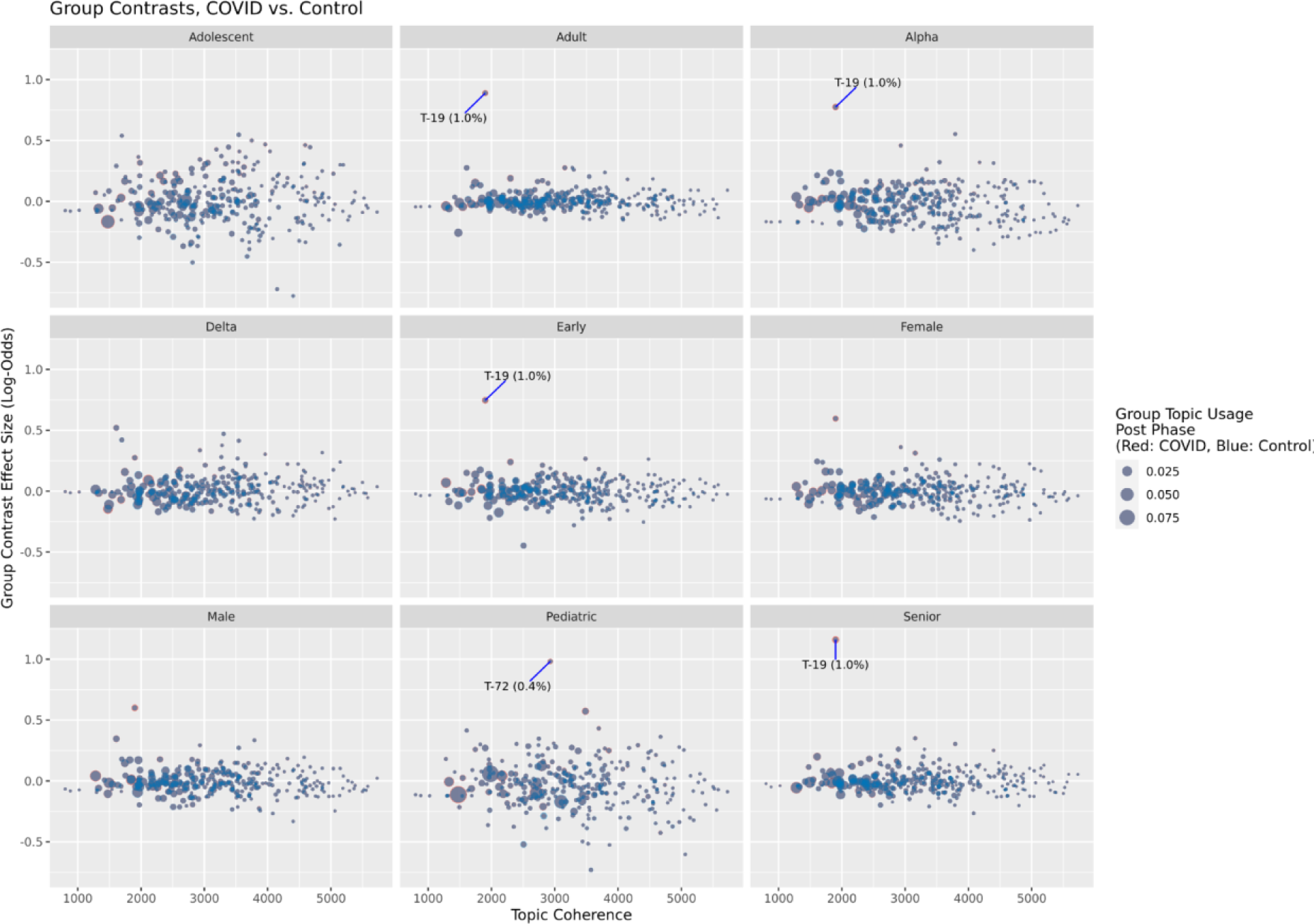
Per-topic coherence (horizontal axis) vs. contrast effect sizes (log-odds scale, vertical axis) for tested groups (panels) in PASC vs. Control (top) and COVID vs. Control (bottom) contrasts. Labeled topics are those with statistically significant log-odds differences of >1 or <-1 (OR >2 or <0.5). Points are sized and colored according to mean topic usage for the group and cohort in the post-infection phase, with blue points representing Control patients and red points representing PASC (top) or COVID (bottom) patients.

**Suppl. Figure 9.**
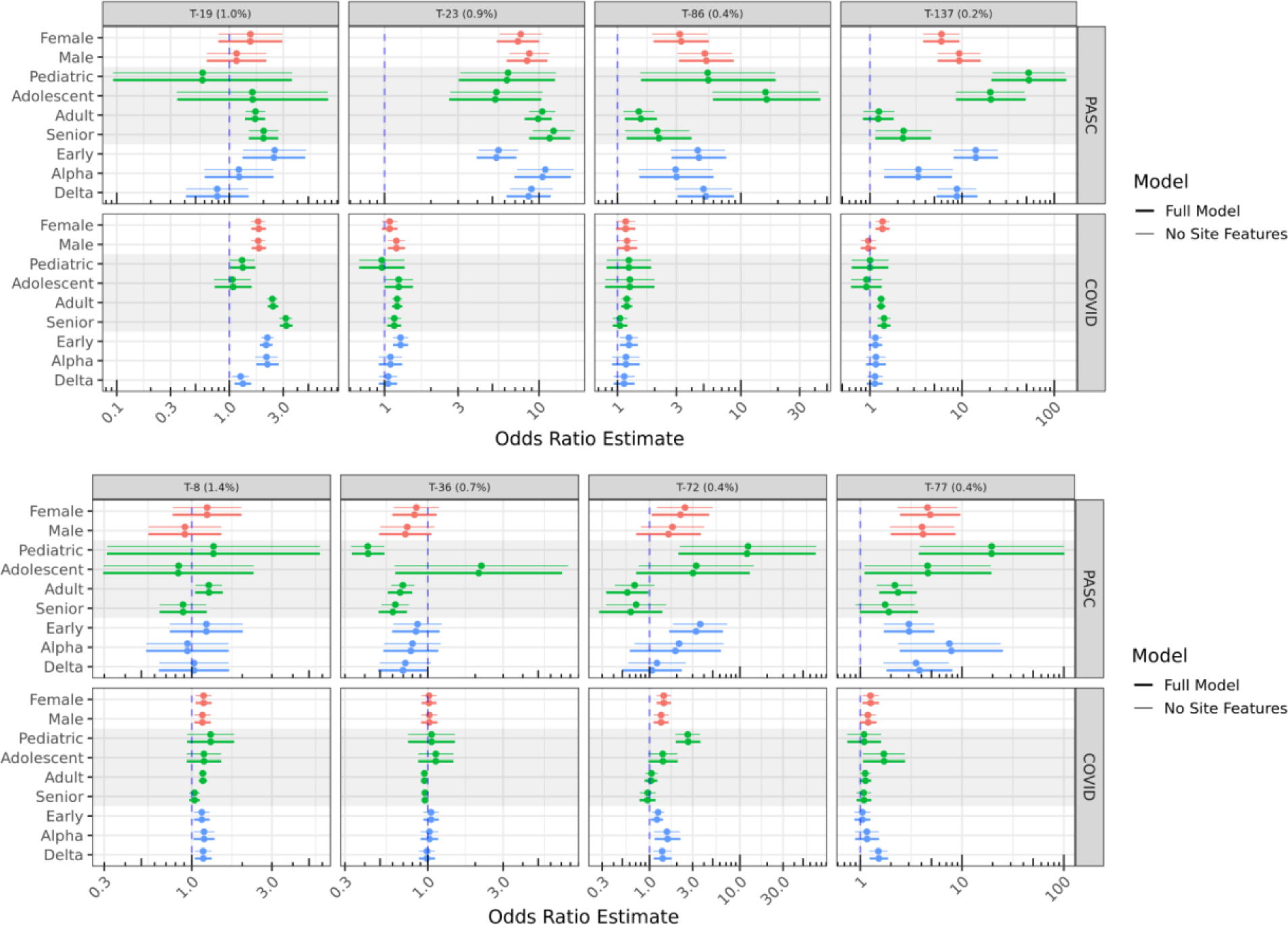
Results for Figures 4 (top) and 5 (bottom) for models with and without site-level covariates of topic usage, percentage of PASC patients, and source common data model.

## Supplemental Methods

### Minimal Site Quality Filters

EHR data from the National COVID Cohort Collaborative (N3C), released Aug. 2, 2022 represent records from 75 contributing sites. All analyses were restricted to data from 63 sites passing minimal quality checks: sites were excluded if greater than 25% of inpatient visits were not accompanied by serum creatinine or white blood cell count measures (N=11), or if greater than 5% of COVID-19 confirmed patients were indicated as inpatient continuously for 200 or more days prior to and including their confirmed COVID-19 date (as potential long-term care facilities, N=1).

### Model Training

Model training utilized the online Latent Dirichlet Allocation (LDA) method of Hoffman et al.^24^ as implemented in Apache Spark (pyspark.ml.clustering.LDA) version 3.2.1.^29^ Parameters used include k (the number of topics, 300 in the final model), seed (42, a random seed to initialize the training), and maxIter (200, providing 10 passes over the training data in batches of 5% each). Determination of condition-topic and topic-patient distributions were produced by the fitted LDA model.

### Topic Annotations

Each topic is annotated with three values: U, representing the relative usage of the topic by total weight assigned to patients (range 0-100%), H, a measure how uniformly the topic is used by N3C-contributing sites (range 0-1, with values closer to 0 being site-specific), and C, a measure of each topics’ coherence compared to the mean over all topics. All three are computed over the training and validation sets.

U is computed as the sum over patients of the weight assigned to the topic, divided by the number of patients (which is also the total weight assigned over all topics).

H is computed as the information entropy of the relative usage of the topic across sites, normalized to a maximum value of 1.0 when the usage is uniformly distributed. Relative usage for a given site is computed as the total weight assigned to the topic for patients from the site, divided by the total number of patients from that site.

Per-topic coherence C is calculated for each topic using the UCI Coherence metric (see Model Validation below). These values are not meant to be interpreted on an absolute scale, but since they are normally distributed amongst topics (Suppl. Figure 4) we adjust them to z-scores for comparative use.

### Jensen-Shannon Distance

Jensen-Shannon Distance between topics *t*_*i*_ and *t*_*j*_ is a true metric and is defined as the square root of the Jensen-Shannon divergence:

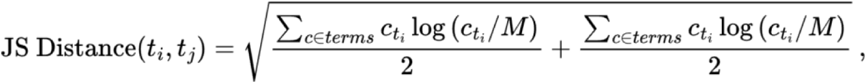

where *c*_*tx*_ = *p*(*c*|*t*_*x*_) (the probability assigned to term *c* in topic *t*_*x*_) and *M* is (*c*_*ti*_ + *c*_*tj*_)/2.

### Topic Term Relevance

Term relevance provides a measure of term-topic-specificity, with values greater than zero indicating terms more likely for the topic than overall.^33^ For term *c*_*i*_ and topic *t*_*j*_, we define relevance as

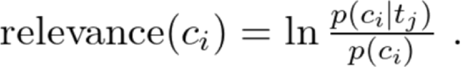

### Model Validation

UCI coherence for a given topic *t*_*i*_ is computed over the top N terms by probability for the topic, where we used N = 20. Letting *T*_*i*_ be the set of top 20 terms for *t*_*i*_, a sum score is computed for each distinct pair of terms a and b, where the score for a given pair is the log of the measured probability of their occurring together in a patient compared to the joint probability assuming independence. To avoid undefined scores, 0 is used for pairs where the denominator is 0, and 1 is added to the joint probability.^30^

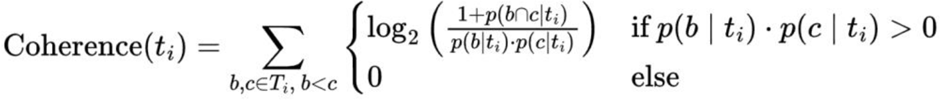

Overall model quality was evaluated as the mean of coherence scores across topics, computed over the validation dataset only.

### Per-Condition Tests

All tests were performed in R v3.5.1.^61^ As described in the main text, patients in the test data set were included for evaluation of new-onset conditions if they satisfied requirements for being in the PASC, COVID, or Control cohorts. The top 20 conditions from each topic with relevance score > 0 were evaluated by considering only patients without the condition in the pre phase, comparing counts of PASC (and COVID) patients later indicated and not indicated for the post phase, to those same counts in the Control cohort. R’s fisher.test() was used with simulate.p.value = TRUE to support tests where counts are large.^34^ Reported p values were multiple-test corrected using Bonferroni’s method.

### BMI and Quan Comorbidity Scores

Patient BMI values used in modeling were the maximum over those reported after Jan. 1 2018, or the maximum of those computed as weight/(height^2^) if no BMI measurement was directly available. Weight values outside 5kg–300kg and height values outside 0.6m–2.43m were excluded from BMI calculations. Quan comorbidity scores^40^ were computed from available source ICD code prefixes as shown in Suppl. Table 7.

### Topic Regression Tests

Regression models were fitted using geepack v1.3.9,^38^ with contrasts computed using emmeans v1.8.9.^62^ Individual patient histories defined by their pre- and post-phase data were assigned topic probability distributions by the fitted LDA model. For each topic, we fitted a logistic regression model with outcome variable being the model-assigned topic probability as the trial success rate with equal weight, from covariates phase (pre or post), cohort (PASC, COVID, or Control), patient life stage and wave of the index date (see main Methods), sex, race, Quan comorbidity score, BMI, source CDM (PCORnet, ACT, OMOP, TrinetX, and OMOP (PedsNet)). To account for potential differential usage of PASC labels or topics, we also included percentage of patients at the given patients’ site in the PASC cohort, and usage of the topic by the patients’ site relative to all sites (summing to 1.0 across sites). Interactions were included for terms of interest for contrasts using the R/geepack formula topic_probability ∼ phase * cohort * (index_wave + sex + life_stage) + site_percent_pasc * phase * cohort + site_relative_topic_usage + race + quan_score + bmi + cdm. Only patients from the assessment set with complete information for all variables were included.

## Supplemental Tables

**Suppl. Table 1.** OMOP Concepts excluded from model training, evaluation, and testing.

| Concept Name | OMOP Concept Id |
| --- | --- |
| No matching concept | 0 |
| Clinical finding | 441840 |
| COVID-19 | 37311061 |
| Viral disease | 440029 |
| Disease due to coronaviridae | 4100065 |
| Sexually abstinent | 764423 |
| Single current sexual partner | 4043045 |
| New sexual partner | 44813701 |
| Sexually active with men | 43021202 |
| Single historical sexual partner | 43021216 |
| Number of current sexual partners - finding | 4276728 |
| Bigamy | 4336540 |
| Sexual activity - two to three times per month | 4012347 |
| Sexual activity - two to three times per week | 4012202 |
| Finding of number of historical sexual partners | 43021214 |
| No longer sexually active | 4043041 |
| Multiple current sexual partners | 4038723 |
| Sexually active with transgender person | 43021204 |
| Number of sexual partners - finding | 4269990 |
| Satisfactory sexual experience | 44811373 |
| Sexual activity - daily | 4012377 |
| Currently not sexually active | 4012376 |
| Never been sexually active | 4145811 |
| Fornication | 4031991 |
| Sexual activity - monthly | 4012348 |
| Sexual activity - weekly | 4012203 |
| Sexual contact with high risk partner | 44789379 |
| Finding of frequency of sexual activity | 4188013 |
| Engages in sexual activity outside marriage | 43021163 |
| Sexually active with women | 43021203 |
| Purposely unmarried and sexually abstinent | 43021238 |
| Sex within a relationship only | 4021660 |
| Sexually active in last month | 37017764 |
| Sexually active | 4043042 |
| Finding relating to sexual activity | 4114865 |
| Sexually active in last year | 37017763 |
| Engages in sexual activity before marriage | 43021162 |
| Sexually active in last six months | 37017762 |
| Multiple historical sexual partners | 43021215 |

**Suppl. Table 2.** OMOP Concepts describing COVID-19 PCR or Antigen tests.

| Concept Name | OMOP Concept Id |
| --- | --- |
| SARS-CoV-2 (COVID-19) N gene [Presence] in Respiratory specimen by Nucleic acid amplification using CDC primer-probe set N2 | 586525 |
| SARS-CoV-2 (COVID-19) RdRp gene [Presence] in Saliva (oral fluid) by NAA with probe detection | 36032174 |
| SARS-related coronavirus RNA [Presence] in Specimen by NAA with probe detection | 723472 |
| SARS-CoV-2 (COVID-19) N gene [Cycle Threshold #] in Specimen by Nucleic acid amplification using CDC primer-probe set N2 | 706155 |
| SARS-CoV-2 (COVID-19) S gene [Cycle Threshold #] in Specimen by NAA with probe detection | 723468 |
| SARS-CoV-2 (COVID-19) N gene [# /volume] (viral load) in Respiratory specimen by NAA with probe detection | 36661370 |
| SARS-CoV-2 (COVID-19) S gene [Cycle Threshold #] in Respiratory specimen by NAA with probe detection | 723467 |
| SARS-CoV-2 (COVID-19) N gene [Presence] in Serum or Plasma by NAA with probe detection | 586520 |
| SARS-CoV-2 (COVID-19) S gene [Presence] in Respiratory specimen by NAA with probe detection | 723465 |
| SARS-CoV-2 (COVID-19) [Presence] in Specimen by Organism specific culture | 586516 |
| SARS-CoV-2 (COVID-19) N gene [Cycle Threshold #] in Specimen by NAA with probe detection | 706167 |
| SARS-CoV-2 (COVID-19) Ag [Presence] in Respiratory specimen by Rapid immunoassay | 723477 |
| SARS-CoV-2 (COVID-19) RNA [Log # /volume] (viral load) in Specimen by NAA with probe detection | 715262 |
| SARS-related coronavirus N gene [Cycle Threshold #] in Specimen by Nucleic acid amplification using CDC primer-probe set N3 | 706172 |
| SARS-CoV-2 (COVID-19) RNA [Presence] in Saliva (oral fluid) by NAA with probe detection | 715260 |
| SARS-CoV-2 (COVID-19) S gene [Presence] in Serum or Plasma by NAA with probe detection | 586519 |
| SARS-CoV-2 (COVID-19) ORF1ab region [Cycle Threshold #] in Respiratory specimen by NAA with probe detection | 723469 |
| SARS-CoV-2 (COVID-19) RNA [Cycle Threshold #] in Specimen by NAA with probe detection | 586529 |
| SARS-related coronavirus E gene [Presence] in Respiratory specimen by NAA with probe detection | 586523 |
| SARS-CoV-2 (COVID-19) ORF1ab region [Presence] in Saliva (oral fluid) by NAA with probe detection | 36031506 |

| Concept Name | OMOP<br>Concept Id |
| --- | --- |
| SARS-CoV-2 (COVID-19) S gene [Presence] in Specimen by NAA with probe detection | 723466 |
| SARS-CoV-2 (COVID-19) RNA [Presence] in Nasopharynx by NAA with non-probe detection | 723476 |
| SARS-CoV-2 (COVID-19) N gene [Presence] in Saliva (oral fluid) by Nucleic acid amplification using CDC primer-probe set N1 | 36032258 |
| SARS-CoV-2 (COVID-19) RNA [Presence] in Nasopharynx by NAA with probe detection | 586526 |
| SARS-related coronavirus E gene [Presence] in Serum or Plasma by NAA with probe detection | 586518 |
| SARS-CoV-2 (COVID-19) S gene [Presence] in Respiratory specimen by Sequencing | 36031213 |
| SARS-CoV-2 (COVID-19) RNA [Presence] in Nose by NAA with probe detection | 757677 |
| SARS-CoV-2 (COVID-19) N gene [Presence] in Specimen by Nucleic acid amplification using CDC primer-probe set N2 | 706154 |
| SARS-CoV-2 (COVID-19) RNA panel - Respiratory specimen by NAA with probe detection | 706158 |
| SARS-CoV-2 (COVID-19) N gene [Presence] in Respiratory specimen by NAA with probe detection | 706161 |
| SARS-CoV-2 (COVID-19) RdRp gene [Cycle Threshold #] in Specimen by NAA with probe detection | 723470 |
| SARS-CoV-2 (COVID-19) RdRp gene [Presence] in Lower respiratory specimen by NAA with probe detection | 36031652 |
| SARS-CoV-2 (COVID-19) N gene [Presence] in Saliva (oral fluid) by NAA with probe detection | 36661378 |
| SARS-related coronavirus+MERS coronavirus RNA [Presence] in Respiratory specimen by NAA with probe detection | 706159 |
| SARS-related coronavirus E gene [Presence] in Specimen by NAA with probe detection | 706174 |
| SARS-CoV-2 (COVID-19) N gene [Presence] in Specimen by Nucleic acid amplification using CDC primer-probe set N1 | 706156 |
| SARS-CoV-2 (COVID-19) RNA [Cycle Threshold #] in Respiratory specimen by NAA with probe detection | 586528 |
| Measurement of Severe acute respiratory syndrome coronavirus 2 antigen | 37310257 |
| SARS-related coronavirus E gene [Cycle Threshold #] in Specimen by NAA with probe detection | 706166 |
| SARS-CoV-2 (COVID-19) Ag [Presence] in Upper respiratory specimen by Immunoassay | 36032419 |
| SARS-CoV-2 (COVID-19) RNA panel - Specimen by NAA with probe detection | 706169 |
| SARS-CoV-2 (COVID-19) RNA [Presence] in Respiratory specimen by NAA with non-probe detection | 36031238 |
| SARS-CoV-2 (COVID-19) RdRp gene [Presence] in Respiratory specimen by NAA with probe detection | 706160 |
| SARS-CoV-2 (COVID-19) N gene [Presence] in Nasopharynx by NAA with probe detection | 715272 |
| SARS-CoV-2 (COVID-19) N gene [Presence] in Nose by NAA with probe detection | 757678 |
| SARS-CoV-2 (COVID-19) RNA [Presence] in Saliva (oral fluid) by Sequencing | 715261 |
| SARS-CoV-2 (COVID-19) RNA [Presence] in Specimen by NAA with probe detection | 706170 |
| SARS-CoV-2 (COVID-19) N gene [Cycle Threshold #] in Specimen by Nucleic acid amplification using CDC primer-probe set N1 | 706157 |
| SARS-CoV-2 (COVID-19) ORF1ab region [Presence] in Respiratory specimen by NAA with probe detection | 723478 |
| SARS-related coronavirus N gene [Presence] in Specimen by Nucleic acid amplification using CDC primer-probe set N3 | 706171 |
| SARS-CoV+SARS-CoV-2 (COVID-19) Ag [Presence] in Respiratory specimen by Rapid immunoassay | 757685 |
| SARS-CoV-2 (COVID-19) RNA [Presence] in Respiratory specimen by Sequencing | 36661377 |
| SARS-CoV-2 (COVID-19) N gene [Log #/volume] (viral load) in Respiratory specimen by NAA with probe detection | 36661371 |
| SARS-CoV-2 (COVID-19) RdRp gene [Cycle Threshold #] in Respiratory specimen by NAA with probe detection | 723471 |
| SARS-CoV-2 (COVID-19) RdRp gene [Presence] in Upper respiratory specimen by NAA with probe detection | 36031453 |
| SARS-CoV-2 (COVID-19) RdRp gene [Presence] in Specimen by NAA with probe detection | 706173 |
| SARS-CoV-2 (COVID-19) N gene [Presence] in Specimen by NAA with probe detection | 706175 |
| SARS-CoV-2 (COVID-19) ORF1ab region [Cycle Threshold #] in Specimen by NAA with probe detection | 706168 |
| SARS-CoV-2 (COVID-19) N gene [Presence] in Respiratory specimen by Nucleic acid amplification using CDC primer-probe set N1 | 586524 |
| SARS-CoV-2 (COVID-19) ORF1ab region [Presence] in Specimen by NAA with probe detection | 723464 |
| SARS-related coronavirus RNA [Presence] in Respiratory specimen by NAA with probe detection | 706165 |
| SARS-CoV-2 (COVID-19) RNA panel - Saliva (oral fluid) by NAA with probe detection | 36032061 |
| SARS-CoV-2 (COVID-19) RNA [Presence] in Respiratory specimen by NAA with probe detection | 706163 |
| SARS-CoV-2 (COVID-19) specific TCRB gene rearrangements [Presence] in Blood by Sequencing | 36031944 |
| SARS-CoV-2 (COVID-19) RNA [Presence] in Serum or Plasma by NAA with probe detection | 723463 |

**Suppl. Table 3.** All indicators of COVID-19 infection (except for PCR and Antigen tests, Suppl. Table 3).

| Concept Name | Concept Id |
| --- | --- |
| SARS-CoV-2 (COVID-19) IgG Ab [Presence] in Serum, Plasma or Blood by Rapid immunoassay | 706181 |
| SARS-CoV-2 (COVID-19) IgA Ab [Units/volume] in Serum or Plasma by Immunoassay | 723459 |
| SARS-CoV-2 (COVID-19) IgM Ab [Presence] in Serum, Plasma or Blood by Rapid immunoassay | 706180 |
| SARS-CoV-2 (COVID-19) IgM Ab [Presence] in DBS by Immunoassay | 36659631 |
| SARS-CoV-2 (COVID-19) IgM Ab [Titer] in Serum or Plasma by Immunofluorescence | 36661373 |
| SARS-CoV-2 (COVID-19) neutralizing antibody [Presence] in Serum by pVNT | 757680 |
| SARS-CoV-2 (COVID-19) IgG+IgM Ab [Presence] in Serum or Plasma by Immunoassay | 723479 |
| SARS-CoV-2 (COVID-19) Ab panel - Serum, Plasma or Blood by Rapid immunoassay | 706176 |
| SARS-CoV-2 (COVID-19) IgG Ab [Titer] in Serum or Plasma by Immunofluorescence | 36661374 |
| SARS-CoV-2 (COVID-19) IgM Ab [Units/volume] in Serum or Plasma by Immunoassay | 706178 |
| SARS-CoV-2 (COVID-19) IgA Ab [Presence] in Serum or Plasma by Immunoassay | 723473 |
| SARS-CoV-2 (COVID-19) neutralizing antibody [Titer] in Serum by pVNT | 757679 |
| SARS-CoV-2 (COVID-19) Ab [Presence] in Serum or Plasma by Immunoassay | 586515 |
| SARS-CoV-2 (COVID-19) IgG Ab [Units/volume] in Serum or Plasma by Immunoassay | 706177 |
| SARS-CoV-2 (COVID-19) S protein RBD neutralizing antibody [Presence] in Serum or Plasma by sVNT | 36031734 |
| SARS-CoV-2 (COVID-19) IgA Ab [Titer] in Serum or Plasma by Immunofluorescence | 36661372 |
| SARS-CoV-2 (COVID-19) Ab [Units/volume] in Serum or Plasma by Immunoassay | 586522 |
| SARS-CoV-2 (COVID-19) IgA+IgM [Presence] in Serum or Plasma by Immunoassay | 757686 |
| Measurement of Severe acute respiratory syndrome coronavirus 2 antibody | 37310258 |
| SARS-CoV-2 (COVID-19) IgG Ab [Presence] in Serum or Plasma by Immunoassay | 723474 |
| SARS-CoV-2 (COVID-19) Ab panel - Serum or Plasma by Immunoassay | 706179 |
| SARS-CoV-2 stimulated gamma interferon [Presence] in Blood | 36031969 |
| SARS-CoV-2 stimulated gamma interferon release by T-cells [Units/volume] in Blood | 36032309 |
| SARS-CoV-2 (COVID-19) IgA Ab [Presence] in Serum, Plasma or Blood by Rapid immunoassay | 586521 |
| SARS-CoV-2 (COVID-19) Ab [Presence] in DBS by Immunoassay | 36031197 |
| SARS-CoV-2 (COVID-19) Ab [Presence] in Serum, Plasma or Blood by Rapid immunoassay | 36661369 |
| SARS-CoV-2 (COVID-19) IgM Ab [Presence] in Serum or Plasma by Immunoassay | 723475 |
| SARS-CoV-2 (COVID-19) Ab [Interpretation] in Serum or Plasma | 723480 |
| SARS-CoV-2 (COVID-19) IgG Ab [Presence] in DBS by Immunoassay | 586527 |
| SARS-CoV-2 stimulated gamma interferon release by T-cells [Units/volume] corrected for background in Blood | 36031956 |

Suppl. Table 4

All significant single-condition tests. Listed estimates are odds ratios for the given cohort pre-to-post compared to Controls, and p-values are adjusted across all condition tests for both cohorts (Bonferroni, prior to filtering to significance). Available at https://doi.org/10.5281/zenodo.11188766.

**Suppl. Table 5.** Summary statistics for patients in the assessment set, with mean and standard deviation of condition era counts in pre- and post-infection phases. Note that the pre-infection phase covers 1 year of patient history, while the post-infection phase covers 6 months post-acute.

| Cohort | Life Stage | Phase | Mean # Conditions | SD # Conditions | # Patients | # Sites |
| --- | --- | --- | --- | --- | --- | --- |
| Control | adolescent | post | 10.296 | 10.373 | 10789 | 32 |
| Control | adolescent | pre | 15.76 | 16.994 | 10789 | 32 |
| Control | adult | post | 17.518 | 18.525 | 180338 | 34 |
| Control | adult | pre | 27.794 | 29.446 | 180338 | 34 |
| Control | pediatric | post | 8.894 | 9.611 | 16029 | 32 |
| Control | pediatric | pre | 15.815 | 19.157 | 16029 | 32 |
| Control | senior | post | 25.357 | 24.142 | 95861 | 33 |
| Control | senior | pre | 40.438 | 36.562 | 95861 | 33 |
| COVID | adolescent | post | 10.311 | 12.376 | 3703 | 31 |
| COVID | adolescent | pre | 15.979 | 19.2 | 3703 | 31 |
| COVID | adult | post | 17.177 | 18.777 | 60279 | 34 |
| COVID | adult | pre | 28.432 | 31.169 | 60279 | 34 |
| COVID | pediatric | post | 10.074 | 12.872 | 3724 | 29 |
| COVID | pediatric | pre | 17.001 | 21.634 | 3724 | 29 |
| COVID | senior | post | 24.847 | 24.162 | 21668 | 34 |
| COVID | senior | pre | 41.522 | 38.15 | 21668 | 34 |
| PASC | adolescent | post | 23.287 | 22.347 | 66 | 20 |
| PASC | adolescent | pre | 18.893 | 21.219 | 66 | 20 |
| PASC | adult | post | 30.281 | 30.778 | 2047 | 32 |
| PASC | adult | pre | 34.566 | 42.242 | 2047 | 32 |
| PASC | pediatric | post | 21.061 | 15.282 | 49 | 18 |
| PASC | pediatric | pre | 19.755 | 15.492 | 49 | 18 |
| PASC | senior | post | 42.374 | 36.527 | 697 | 32 |
| PASC | senior | pre | 50.292 | 51.6 | 697 | 32 |

Suppl. Table 6

All topic-level logistic model tests. Estimates are odds ratios for the given cohort and demographic compared to Controls for the same demographic. Ratios where the demographic is listed as NA are for demographic contrasts independent of phase or cohort (model effectiveness checks, see main Methods). P-values are adjusted across all contrast tests (Holm). Available at https://doi.org/10.5281/zenodo.11188766.

**Suppl. Table 7.** Source ICD code prefixes used to generate Quan-based comorbidity scores.

| ICD Prefixes | Charleson Group | Quan Score |
| --- | --- | --- |
| 'I21','I22','I252' | 1: Acute or historical MI | 0 |
| 'I43','I50','I099','I110','I130','I132','I255','I420','I425','I426','I427','I428','I429','P290' | 2: CHF | 2 |
| 'I70','I71','I731','I738','I739','I771','I790','I792','K551','K558','K559','Z958','Z959' | 3: Peripheral vascular disease | 0 |
| 'G45','G46','I60','I61','I62','I63','I64','I65','I66','I67','I68','I69','H340' | 4: Cerebrovascular disease | 0 |
| 'F00','F01','F02','F03','G30','F051','G311' | 5: Dementia | 2 |
| 'J40','J41','J42','J43','J44','J45','J46','J47','J60','J61','J62','J63','J64','J65','J66','J67','I278','I279','J684','J701','J703' | 6: COPD | 1 |
| 'M32','M33','M34','M06','M05','M315','M351','M353','M360' | 7: Rheumatic disease | 1 |
| 'K25','K26','K27','K28' | 8: Peptic ulcer | 0 |
| 'B18','K73','K74','K700','K701','K702','K703','K709','K717','K713','K714','K715','K760','K762','K763','K764','K768','K769','Z944' | 9: Mild liver disease | 2 |
| 'E100','E101','E106','E108','E109','E110','E111','E116','E118','E119','E120','E121','E126','E128','E129','E130','E131','E136','E138','E139','E140','E141','E146','E148','E149' | 10: Diabetes | 0 |
| 'E102','E103','E104','E105','E107','E112','E113','E114','E115','E117','E122','E123','E124','E125','E127','E132','E133','E134','E135','E137','E142','E143','E144','E145','E147' | 11: Diabetes with chronic complications | 1 |
| 'G81','G82','G041','G114','G801','G802','G830','G831','G832','G833','G834','G839' | 12: Paralysis | 2 |
| 'N18','N19','N052','N053','N054','N055','N056','N057','N250','I120','I131','N032','N033','N034','N035','N036','N037','Z490','Z491','Z492','Z940','Z992' | 13: Renal disease | 1 |
| 'C00','C01','C02','C03','C04','C05','C06','C07','C08','C09','C10','C11','C12','C13','C14','C15','C16','C17','C18','C19','C20','C21','C22','C23','C24','C25','C26','C30','C31','C32','C33','C34','C37','C38','C39','C40','C41','C43','C45','C46','C47','C48','C49','C50','C51','C52','C53','C54','C55','C56','C57','C58','C60','C61','C62','C63','C64','C65','C66','C67','C68','C69','C70','C71','C72','C73','C74','C75','C76','C81','C82','C83','C84','C85','C88','C90','C91','C92','C93','C94','C95','C96','C97' | 14: Localized cancer/leukemia/lymphoma | 2 |
| 'K704','K711','K721','K729','K765','K766','K767','I850','I859','I864','I982' | 15: Moderate/severe liver disease | 4 |
| 'C77','C78','C79','C80' | 16: Metastatic cancer | 6 |
| 'B20','B21','B22','B24' | 17: HIV/AIDS | 4 |

